# Tandem Repeat Polymorphisms Are Associated with Brain Structure: Results of Two Large Population-based Studies

**DOI:** 10.1101/2025.11.14.25340228

**Authors:** Richard Mantey, Jialu Hu, Maryam Touhidinia, Tanzeem Butt, Ahmed M. Sidky, Roohollah Sobhani, Santiago Estrada, Kristian Haendler, Elena De Domenico, Marc D. Beyer, Monique M.B. Breteler, N. Ahmad Aziz

**Author notes:** Corresponding Author: Prof. Dr. N. Ahmad Aziz, MD PhD Population & Clinical Neuroepidemiology, German Center for Neurodegenerative Diseases (DZNE) Venusberg-Campus 1/99, 53127 Bonn Germany.

## Abstract

Although genome-wide association studies (GWAS) have uncovered many genetic variants linked to brain structure, much of its heritability remains unexplained. Short tandem repeats (STRs), which are rarely considered in GWAS, may contribute to this “missing heritability”. Using targeted deep sequencing, we systematically assessed the relationship between ∼3000 polymorphic STRs and brain imaging-derived phenotypes across 2958 participants of the Rhineland Study. Expansion of an intronic CA repeat in *PRR14L* was associated with larger thalamic volume (standardized β [95% CI]=0.15 [0.06–0.24]), while AATG repeat polymorphisms in *NADK* were associated with reduced subcortical gray matter volume (–0.05 [–0.08 to –0.01]). Both associations replicated in the UK Biobank cohort (N=38879). Beyond single loci, higher polygenic burden of moderate STR expansions was associated with increased total brain, gray matter, supratentorial, and thalamic volumes. Our findings indicate that moderate STR expansions are region-specific determinants of brain morphology and suggest that STR variability may have evolved to enhance neuroanatomical plasticity and cognitive function.

**Teaser:** Hidden DNA repeats shed light on the genetics behind brain diversity.

## Introduction

A large proportion of the risk for common age-associated brain disorders, such as Alzheimer’s and Parkinson’s disease, is genetically determined. Over the past few decades, genome-wide association studies (GWAS) have provided insights into the genetic underpinnings of many neurodegenerative and age-related brain diseases (*1–4*). However, a substantial amount of the genetic basis of these diseases still remains unknown. A potential cause of this “missing heritability” conundrum is that many GWAS have predominantly focused on the role of single nucleotide polymorphisms (SNPs), often neglecting the potentially important contributions of other sources of genomic variation. Repetitive regions of the human genome such as retrotransposable elements and short tandem repeats (STRs) are among the most polymorphic structural genetic variants and may account for part of the “missing heritability” problem of brain disorders.

STRs, also referred to as microsatellites, are generally defined as DNA sequences consisting of repeated motifs of 1-6 base pairs (*5*). These conspicuous sequences are not only ubiquitous in the human genome but are also substantially more polymorphic than SNPs due to replication slippages (*6*). Variations in STRs copy numbers therefore contribute substantially to genetic diversity (*7*). STRs are particularly known for their genomic instability, which often leads to dynamic changes in repeat length across generations as well as within somatic tissues (*8*). Indeed, genomic instability-induced repeat expansion in certain genes is thought to have given rise to repeat expansion disorders (REDs), most of which are characterized by neurological abnormalities (*8*). Even though the pathological consequences of large repeat expansions are well documented, much less is known about the potential effects of repeat length variation within the normal or premutation range of the respective loci. This knowledge gap is particularly striking for brain-related phenotypes in the general population. STRs are prone to somatic expansion over time due to replication-associated mutations, which suggests that even subtle variations in repeat length could influence brain structure and function across the lifespan (*9*). Understanding these dynamics could therefore provide valuable insights into the broader role of STR variations as determinants of brain structure, particularly in the context of aging.

An important implication of age-related somatic instability is the possibility that STR variants exert age-dependent effects, particularly on cognition and other brain-related traits. We previously demonstrated that cytosine-adenine-guanine (CAG) repeat length variations in three polyglutamine disease-associated genes (PDAGs) were significantly associated with cognitive decline (*10*). Furthermore, we observed that large normal-range CAG repeats in *TBP* and *ATXN7* genes were linked to an increased lifetime risk of depression (*11*), while those in *AR* and *ATXN1* gene modified disease expression in Alzheimer’s disease (*12*). Collectively, these findings underscore the potential role of STRs in modulating brain function even below the pathogenic expansion threshold associated with REDs.

The direction and magnitude of the effects of non-and sub-pathological allele lengths in REDs genes, however, remain to be fully elucidated. Interestingly, the Kids-HD study, a longitudinal study of early manifestations of Huntington’s disease (HD) in children and young adults, demonstrated that CAG repeat mutation carriers in the *HTT* gene exhibited significantly better cognitive, behavioral, and motor performance than non-expanded gene carriers in early life. The characteristic structural and functional brain decline emerged later, following this initial peak in brain performance (*13*). These results align with the evolutionary theory of antagonistic pleiotropy, which was first proposed by Williams in 1957 and has been refined and supported by recent findings from other species (*14*). This theory posits that certain genetic variants that may confer advantages early in life could become detrimental in later years (*15*). This may be particularly the case for some human-specific STR loci that initially may have evolved to confer advantages in cognitive capacity (*16–18*).

The examples detailed above represent only a minute fraction of the vast STR landscape. The human genome contains hundreds of thousands of STRs whose potential influence on brain-related traits remains largely unexplored. Therefore, here we aimed to systematically investigate the relationship between STR polymorphisms and brain imaging-derived phenotypes across the adult lifespan in two large, well-characterized, population-based cohorts of European ancestry. Our findings indicate that moderate STR expansions, particularly in genes linked to neurodegeneration, can indeed act as region-specific determinants of brain morphology and provide further support for the antagonistic pleiotropy hypothesis of STR expansions in humans.

## Results

### STR profiling in two independent cohorts

For discovery we utilized data from the first 2,958 participants of the population-based Rhineland Study in whom both brain imaging data and DNA samples were available (see **Table 1** for participant characteristics). We performed targeted sequencing (with paired-end 2 × 250 bp reads) of ∼3,000 STR loci using an amplicon-based targeted sequencing approach on the Illumina platform. For replication of our findings, we used the UK Biobank (UKB) as an independent cohort. Specifically, we focused on participants with available brain imaging data from the UKB imaging sub-study (*19*). Of the ∼50,000 individuals with imaging data available, whole genome sequencing (WGS) data were available for 45,306 participants. WGS in UKB was also conducted using the Illumina platform with ∼30× coverage and paired-end 2 × 75 bp reads (*20*). Unlike the Rhineland Study, which is a relatively homogeneous cohort of predominantly European ancestry, the UKB is more genetically diverse. Therefore, to minimize population structure differences between the cohorts, we restricted our replication analyses to 38,879 UKB participants of genetically determined British ancestry (**Table 1**).

**Table 1:** Participant characteristics in the Rhineland Study and UK Biobank Imaging Substudy.

| Variable | RS Cohort | UKB Cohort |
| --- | --- | --- |
| Sample size (n) | 2958 | 38879 |
| Age, mean (SD) | 54.1 (13.5) | 55.1 (7.5) |
| Age range (years) | 30-90 | 40-70 |
| Female, n (%) | 1688 (57.1 %) | 20518 (52.8 %) |

**Abbreviations:** RS: Rhineland Study, SD: standard deviation, UKB: UK Biobank

### Population characteristics

Among participants included in the Rhineland Study, the mean age was 54.1 ± 13.5 years, with women comprising 57.1% of the sample. On the other hand, participants from the UKB study had a mean age of 55.1 ± 7.5 years, with 52.8% women. Additional characteristics of the brain imaging-derived phenotypes such as global measures (e.g., total brain volume, total gray matter volume) and subcortical structures (e.g., thalamus and related regions) are provided in **Supplementary Table S1**.

### Selection and characterization of STR loci for association testing

To investigate the effect of STR polymorphisms on brain health, we first conducted comprehensive targeted genetic association analyses. Our initial customized STR catalogue included 2,817 loci which were carefully curated based on their previously reported roles in brain structure and function, as well as their potential association with neurological disorders as identified through comparative genomic studies(*17, 18, 21–24*). Additionally, highly polymorphic STR loci commonly used in DNA fingerprinting and linkage studies – such as those included in the Marshfield genetic map – were included in our catalogue (*25*). From this, we focused on 2,447 STRs that were polymorphic, defined here as showing variability in tract length across the participants of the Rhineland Study. The genomic distribution of these analysed STRs is summarized in **Fig. 1**.

**Fig. 1:**
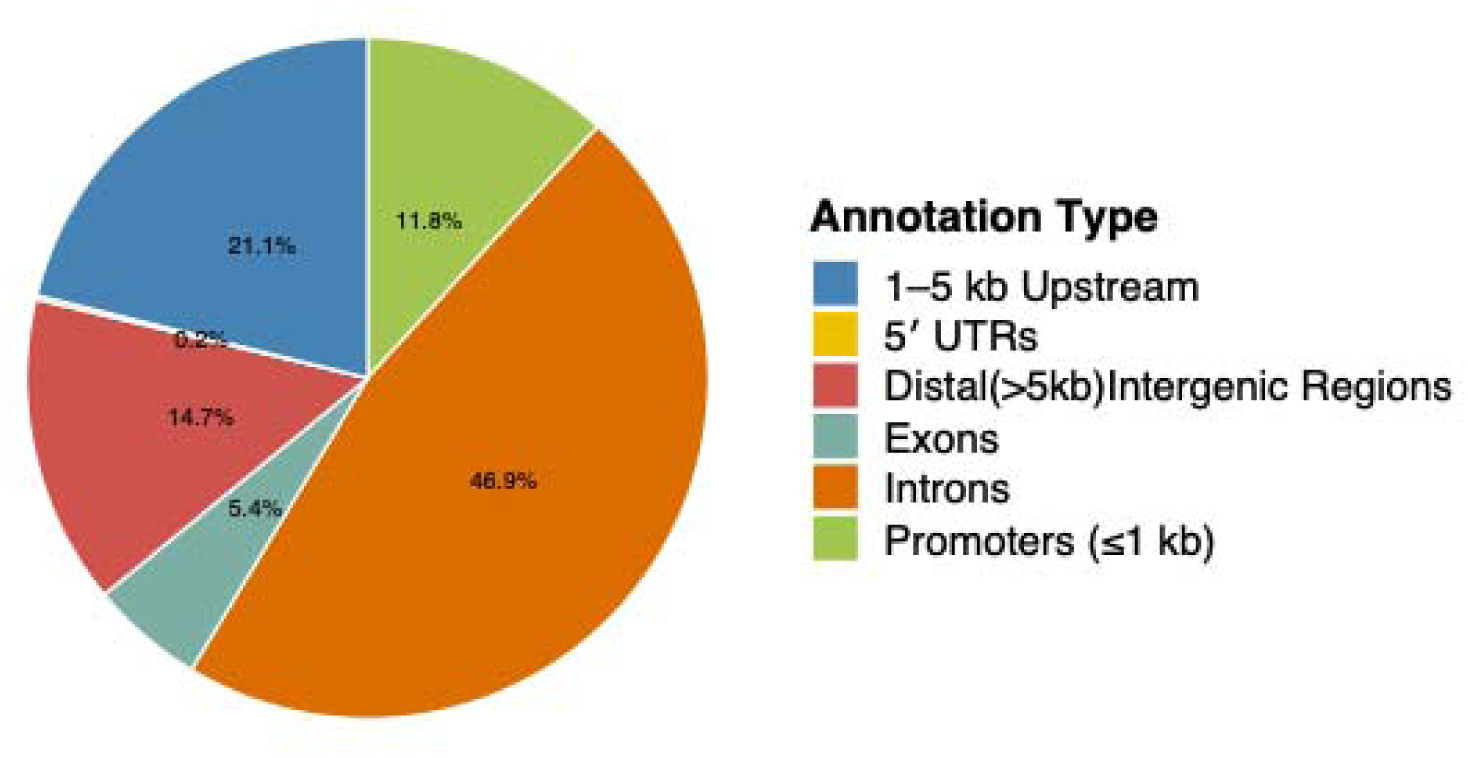
Genomic distribution of STRs in our customized STR catalogue. This figure illustrates the genomic distribution of STR loci included in our customized STR panel. The majority of STRs (46.9%) were located within intronic regions, followed by distal intergenic regions (21.1%), which were defined as sequences outside gene bodies and at least 3 kb from the nearest transcription start site (TSS). Additionally, 11.8% of the loci were positioned 1–5 kb upstream of the TSS, while 14.7% reside within promoter regions (defined as <1 kb upstream of the TSS). Only a small proportion of loci were located within exonic (5.4%) or untranslated regions (UTRs, 0.2%). **Abbreviations:** UTR, untranslated region; TSS, transcription start site

### Locus-specific genetic association analyses

In our genetic association analyses, we used allelic dosage, defined here as the mean repeat length of the two alleles per STR locus, as the primary independent variable in a multiple linear regression framework. We then applied a rank-based inverse-normal transformation to account for the non-normal distribution observed at most STR loci. All models were adjusted for age, sex, the first 10 genetic principal components as well as other relevant covariates as described in the ‘Statistical Analyses’ section of the **Methods**. In the Rhineland Study discovery cohort, we observed heterogeneous patterns of association between STR loci and brain morphology across multiple regions. While some loci were related to global volumetric measurements, others exhibited a more region-specific effect. Thalamic volume showed the strongest enrichment, with 31 significant associations, followed by total gray matter volume (30 loci) and total brain volume (15 loci) (**Fig. 2**). Additional associations were detected for supratentorial (7 loci) and subcortical gray matter volumes (8 loci), with several loci overlapping across brain regions (**Fig. 2**).

**Fig. 2:**
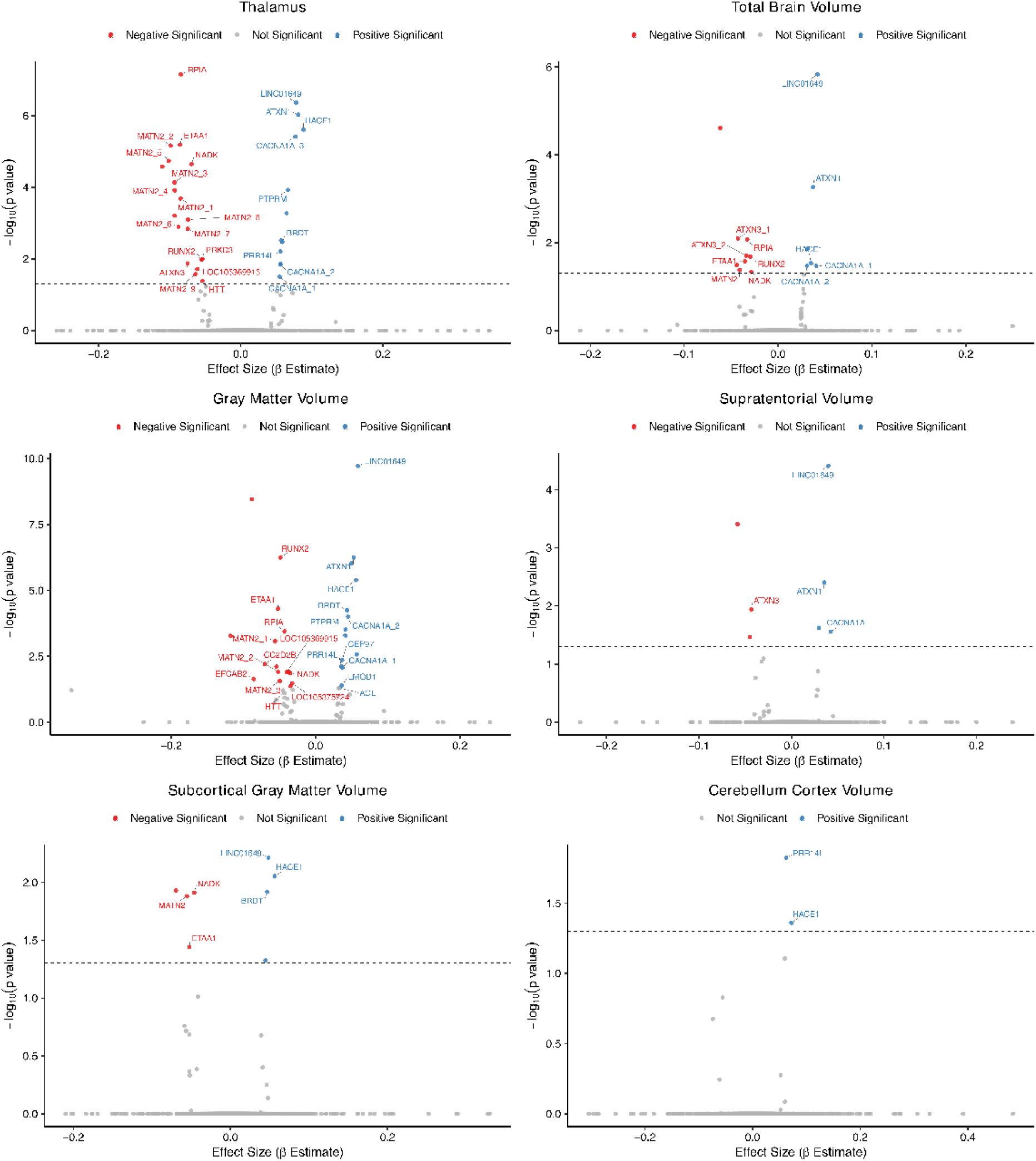
Associations between STR loci and regional brain volumes. Volcano plots depict the associations between STR loci and regional brain volumes. The x-axis represents the effect size (β estimates) and the y-axis the –log10 p-values. Black dashed lines indicate the FDR significance thresholds. Genes annotated in blue are positively associated with the respective brain imaging-derived phenotype, whereas those in red are negatively associated. Gene suffix numbers indicate distinct loci within the same gene to which short tandem repeats were mapped in the gene proximity analysis. A heterogeneous pattern of associations was observed across brain regions. Notably, 31 loci were significantly associated with thalamic volume, 30 with total gray matter volume, and 15 with TBV. Additional associations were identified for supratentorial gray matter (7 loci) and subcortical gray matter volumes (8 loci), with several loci overlapping across brain regions.

Interestingly, some of the overlapping genes include both well-established disease-causing loci and novel candidates. For instance, extreme repeat expansions in some of these overlapping genes such as *ATXN1* and *CACNA1A* are known to cause neurodegenerative diseases in humans, whereas repeat length polymorphisms in others, including *PRR14L*, *RUNX2*, and *MATN2*, have not been previously implicated in neurological disorders. These findings highlight both known disease-associated STRs and novel candidates that may play broader roles in influencing brain structure.

To externally validate the identified associations, we tested the STRs that retained statistical significance after false discovery rate (FDR) correction in the Rhineland Study for association testing with the respective brain imaging-derived phenotypes in the UKB cohort. We applied a Bonferroni-adjustment threshold (α = 0.05/n, where *n* is the number of loci carried forward for replication). STR loci with *p*-values less than this threshold in the UKB were considered statistically replicated. In our replication analysis, more than half (60.2%) of the STR loci tested for the different brain imaging-derived phenotypes showed consistent directions of effect between the discovery and replication cohorts. Among these, about 30% of the loci reached nominal significance, i.e. *p* < 0.05 (**Supplementary Fig. S2**). Using the predefined Bonferroni-adjusted significance threshold, we identified robust replication signals for both thalamic and subcortical gray matter volumes. Specifically, CA repeat motif variations at chr22, located in the intronic region of the *PRR14L* gene, were significantly associated with larger thalamic volume (β = 0.15, 95% CI [0.06 to 0.24]) (**Fig. 3)**. In addition, polymorphisms of an AATG repeat motif in *NADK* were significantly associated with reduced subcortical gray matter volume (β = –0.05, 95% CI [–0.08 to –0.01]). Thus, both loci demonstrated consistent replication in the UK Biobank which may highlight their importance in brain morphology. A comprehensive list of all tested loci, including those that did not meet statistical thresholds for replication, is provided in **Supplementary Fig. S2**.

**Fig. 3:**
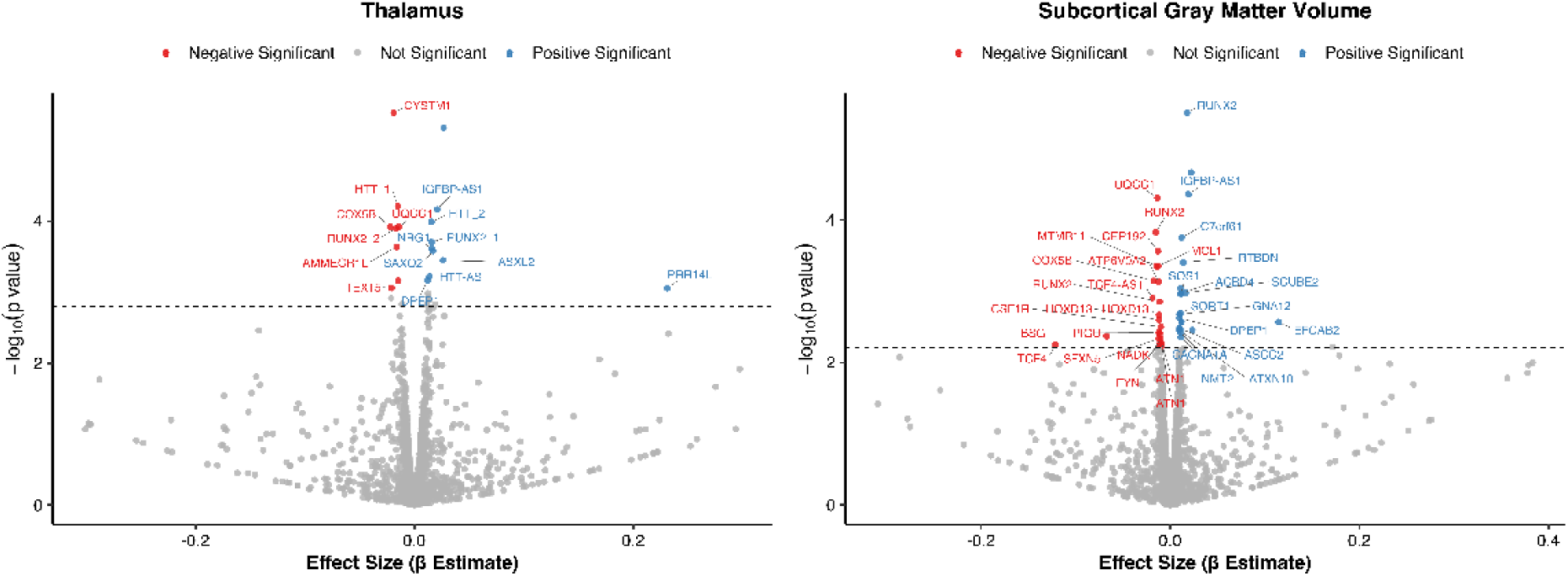
Replication of STR associations in the UK Biobank cohort. Volcano plots for each brain region show estimates of STR loci associations in the replication analysis. While many loci demonstrated consistent directions of effect with nominal significance (p < 0.05), two surpassed the FDR-corrected threshold (adjusted p < 0.05). Specifically, repeat polymorphism in *PRR14L* was significantly associated with increased thalamic volume (β = 0.15, 95% CI [0.06 to 0.24]), and an AATG repeat expansion near *NADK* was significantly associated with reduced subcortical gray matter volume (β = –0.05, 95% CI [–0.08 to –0.01]).

### Aggregated STR length and brain morphology

To assess the association of STR lengths aggregated over all the genotyped loci with brain structure, we z-standardized the mean allele size at each locus and summed these values to derive an aggregate standardized mean allele score. This approach enabled capturing the cumulative variation in STR length across the target loci while reducing locus-specific scaling effects. The aggregated STR length was then included as the independent variable in the regression models with brain imaging-derived phenotypes as the dependent variables.

We observed that a higher aggregated STR length was associated with increased overall brain volume. While the association with total brain volume did not reach statistical significance in the Rhineland Study, it was significant in the UKB. Conversely, a larger aggregated STR length was significantly associated with increased thalamic volume in the Rhineland Study but did not reach statistical significance in the UKB (**Supplementary Fig. S3**). Importantly, however, the highly consistent direction and magnitude of effects across multiple brain regions and across both cohorts support a potential biological role of STR length variations in shaping brain morphology.

### Polygenic STR burden and brain morphology

In our primary genetic association analysis, we modeled STR tract lengths as continuous variables in relation to the brain imaging-derived phenotypes. We used this approach in order to capture the full spectrum of naturally occurring STR variation and its potential contribution to inter-individual differences in brain structure. Moreover, an intriguing aspect of allele length polymorphisms is understanding the influence of allele sizes that have not yet reached pathological thresholds on brain phenotypes. While pathogenic STR expansions are typically defined by well-established length thresholds associated with disease onset and severity, it remains unclear whether non-expanded or moderately expanded alleles contribute to the normal variation in brain morphology. Among the 2,817 STR loci included in our customized catalogue, fewer than 100 have known pathogenic thresholds. Therefore, rather than applying arbitrary cutoffs, we developed a STR polygenic burden as previously described by Guo and colleagues (*26*). In this framework, we used pre-specified expansion thresholds relative to the GRCh38 reference genome (≥1, ≥5, ≥10, or ≥20 repeat units) for each locus. While these thresholds are not inherently pathogenic, they allowed us to evaluate whether associations were primarily driven by common alleles or by more extreme expansions in the general population.

In both cohorts, as the expansion threshold increased, the number of STR expansions per individual decreased, highlighting the relative rarity of large expansions in the population (**Supplementary Fig. S4**). In the discovery cohort, we identified significant associations between polygenic STR burden and increased brain volumes across multiple global and regional measures. After correcting for multiple testing using the Li and Ji method (*27*), we found that individuals with a higher polygenic STR burden (≥5 repeat units longer than the reference sequence) exhibited significantly larger volumes in key brain regions, including the thalamus (β = 0.05, 95% CI [0.02 to 0.07]), total brain; β = 0.02, 95% CI [0.007 to 0.03]), and total gray matter (β = 0.03, 95% CI [0.01 to 0.04]) (**Fig. 4**). These associations remained statistically significant also at additional thresholds (≥10 and ≥20 repeats) (**Fig. 5**). Further associations were observed for supratentorial volume, subcortical gray matter volume, and cerebellar cortex volume at the ≥10 and ≥20 repeat thresholds. At the regional level, polygenic STR burden (≥10 repeats) was significantly associated with larger cortical thickness in the left rostral anterior cingulate cortex and left medial orbitofrontal cortex **(Supplementary Fig. S5**). These effects were consistent and remained statistically significant after correction for multiple testing.

**Fig. 4.**
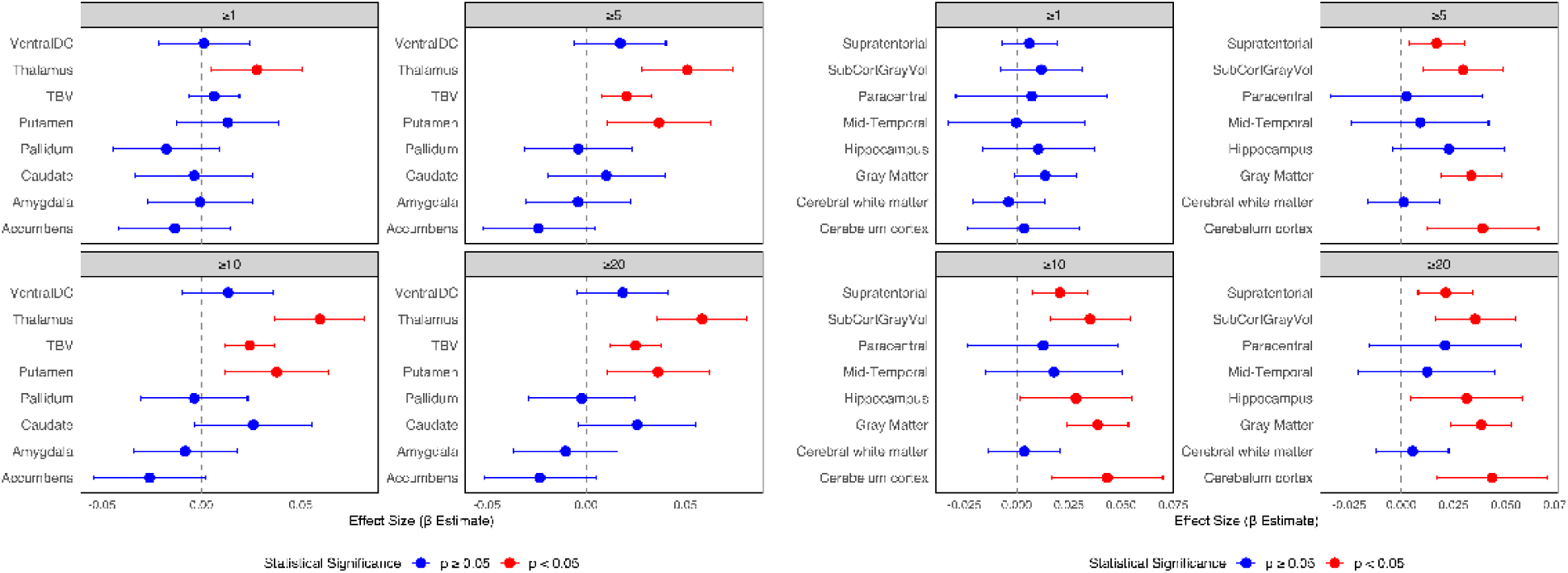
Polygenic STR expansion burden and brain phenotypes in the Rhineland Study. Forest plot of standardized regression coefficients (β) with 95% confidence intervals for the association between polygenic STR expansion burden and brain phenotypes in the Rhineland Study discovery cohort. Each point corresponds to the standardized β estimate for a given brain region at a specific STR threshold and the horizontal bars indicate 95% confidence intervals. Associations remaining after correction for multiple testing using the Li & Ji method are shown in red and non-significant associations are shown in blue. The vertical dashed line represents the null effect (β = 0). **Abbreviations:** TBV, total brain volume; ventral DC, ventral diencephalon

**Fig. 5.**
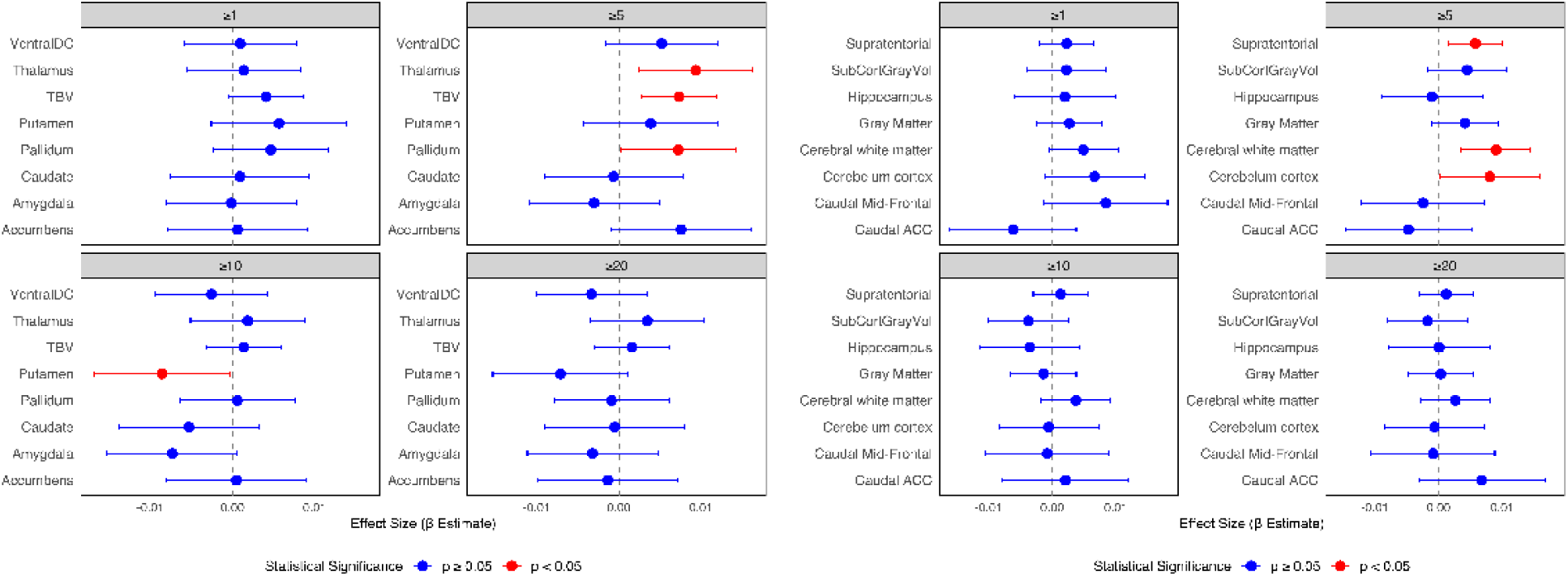
Polygenic STR expansion burden and brain phenotypes in the UKB. Forest plot of standardized β with 95% confidence intervals for the association between polygenic STR expansion burden and brain phenotypes in the UK Biobank replication cohort. Each point corresponds to the β estimate for a given brain region at a specific STR threshold, with horizontal bars indicating the 95% CI. Associations that remained after correction for multiple testing using the Li & Ji are shown in red whereas non-significant associations are shown in blue. The vertical dashed line represents the null effect (β = 0). Statistically significant replicated associations were observed for TBV, thalamic volume, and supratentorial volume for STR expansion burden defined as ≥5 repeat units above the reference. **Abbreviations:** TBV, total brain volume; ventral DC, ventral diencephalon; caudal ACC, caudal anterior cingulate cortex.

We sought to replicate the associations reported above between the polygenic STR expansion burden and brain phenotypes in the UKB cohort. Consistent with our discovery results, we found that individuals with a higher polygenic STR expansion burden (≥5 repeat units longer than the reference sequence) exhibited significantly larger brain volumes. Specifically, we replicated significant positive associations for total brain volume; β = 0.007, 95% CI [0.003 to 0.011] and thalamic volume (β = 0.009, 95% CI [0.002 to 0.016]) (**Fig. 5**). For the remaining brain imaging-based phenotypes tested, the directions of effect were consistent with those observed in the discovery cohort, with the majority reaching nominal significance (p < 0.05) (**Supplementary Fig. S6**).

### Age-dependent effects of polygenic STR burden on brain volume

Given that STRs are prone to genomic instability, which can lead to repeat expansions with increasing age, we examined the age-dependent effects of polygenic STR burden on brain volume, focusing on the associations that replicated. Participants were grouped into 10-year age bins within each cohort, and we assessed the pattern of effect sizes for the ≥5 repeat expansion threshold across these age groups. Interestingly, the relationship between polygenic STR burden and both total brain volume and thalamic volume followed a non-linear trend, with the strongest effects observed between ages 60 and 70 (**Fig. 6**). In the Rhineland Study, the effect size declined after age 70, although this change was not statistically significant, while in the UKB cohort, this could not be evaluated due to the upper age limit of participants.

**Fig. 6:**
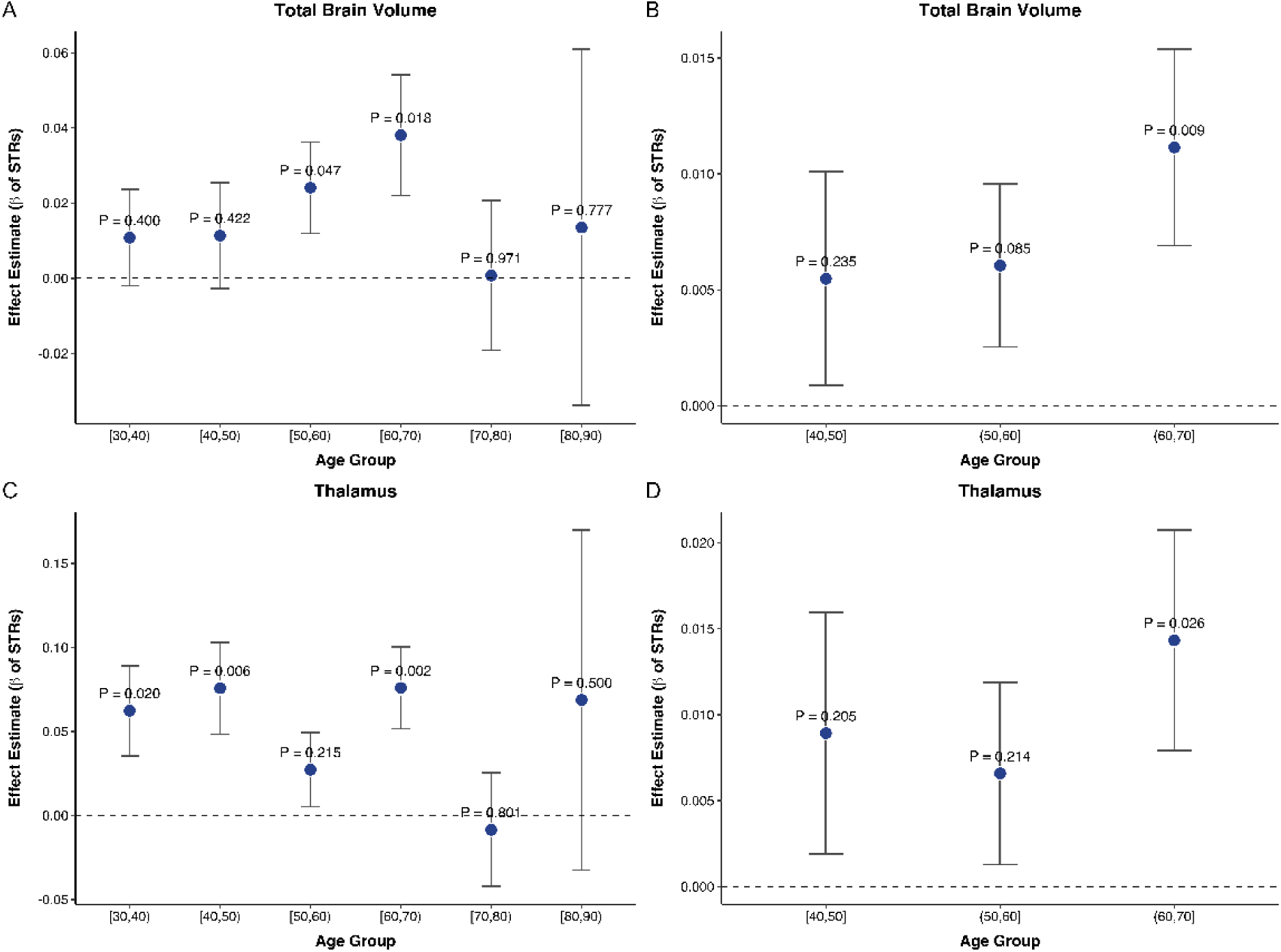
**Age-stratified effects of STR expansion burden on TBV and thalamic volume**. Results are shown for the Rhineland Study (A and C) and UKB (B and D) panels. The strongest associations were observed in participants aged 60–70 years in both cohorts. In Rhineland Study, the effect for TBV reversed direction in the 70–80 age group, even though not statistically significant. These patterns suggest a possible age-dependent effect, with the most pronounced associations in mid-to-late adulthood.

### Pathway enrichment of genes with moderately expanded STR

To better understand the biological mechanisms mediating the relation between STRs and brain structure, we carried out gene ontology (GO) and pathway enrichment analyses at different levels of repeat expansion thresholds. Prior to this, we conducted a genome-wide annotation of approximately 1.2 million STRs identified in the GRCh38 human reference genome (*28*). We then performed a proximity search against annotated human genes from the UCSC Genome Browser (GRCh38 reference genome) to identify the nearest gene for each STR locus. We considered STRs located within a maximum distance of 250 kb from a gene’s transcription start site (TSS) for further analysis. We next ranked the genes in our customized STR panel at predefined expansion thresholds based on carrier frequency within each cohort. At each threshold (≥1, ≥5, ≥10, or ≥20 repeat units longer than the reference), we identified the most frequent genes shared between cohorts and used these for gene set enrichment analysis.

In our enrichment analysis, we identified sets of enrichment terms that were consistent across the moderate expansion thresholds. At the ≥5 repeat units’ threshold, the common top enriched terms were genetic anticipation, spinocerebellar ataxia, progressive cerebellar ataxia, impaired smooth pursuit, and fasciculations, which suggest convergence on neurological features (**Fig. 7**). At ≥10 units threshold, the enrichment terms genetic anticipation and progressive cerebellar ataxia remained and expanded to include gaze-evoked nystagmus and fasciculations. At the highest threshold of ≥20 units, however, the overlap shifted toward molecular and cellular processes. The enrichment terms included TFIIB-class transcription factors, RNA polymerase II transcription initiation, and development of pulmonary dendritic cells and macrophages, alongside neurological terms such as spinocerebellar ataxia (**Fig. 7**).

**Fig. 7:**
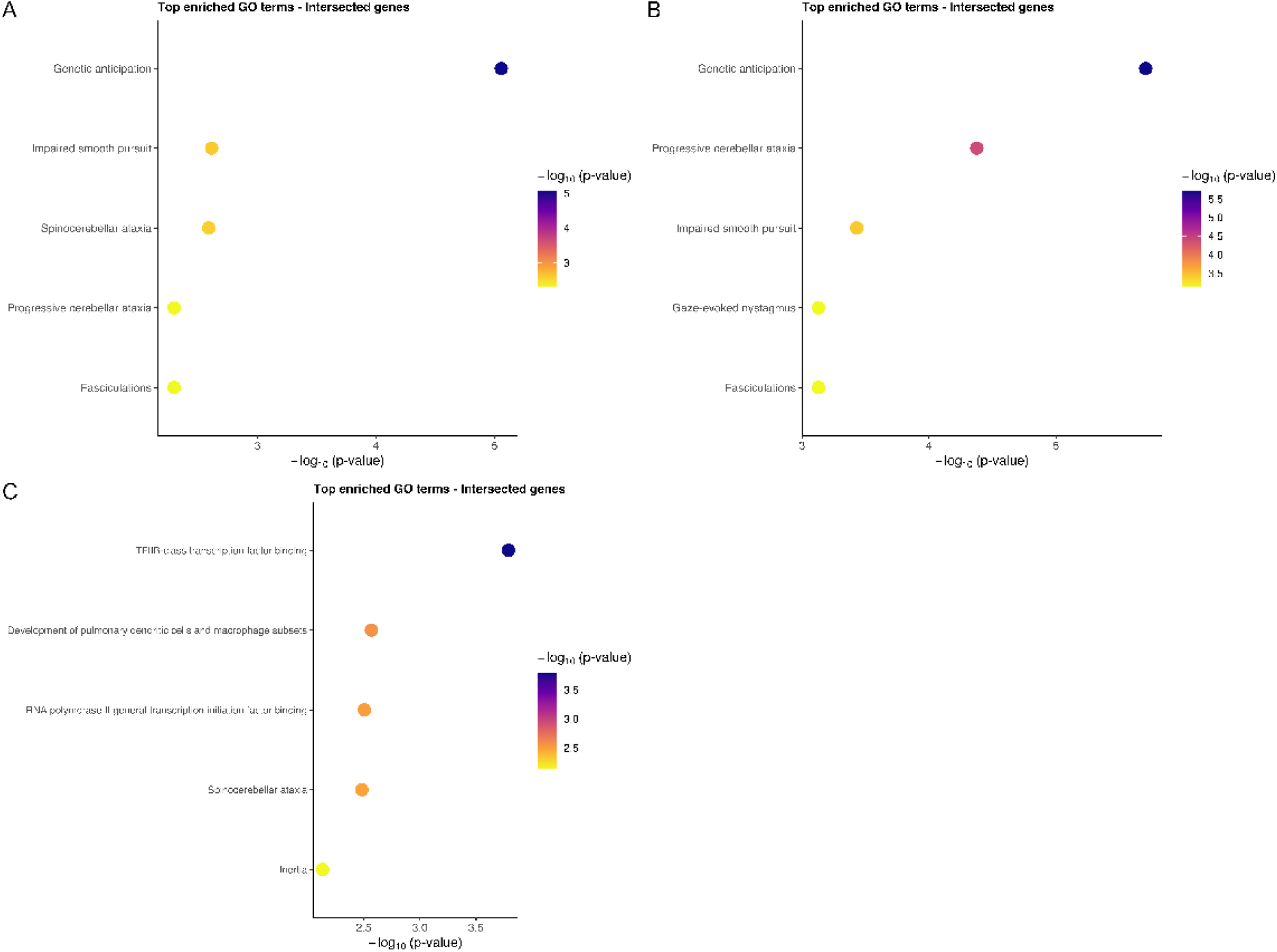
Pathway enrichment results across STR expansion thresholds. Pathway enrichment analysis of genes linked to STR expansions at ≥5, ≥10, and ≥20 repeat units threshold revealed both shared and threshold-specific biological themes. A and B: At ≥5 and ≥10 units threshold, the top enriched pathways were largely neurological (including genetic anticipation, progressive cerebellar ataxia, spinocerebellar ataxia, impaired smooth pursuit, gaze-evoked nystagmus and fasciculations). C: At the ≥20-unit threshold, enrichment shifted toward transcriptional and immune-related processes such as *TFIIB-*class transcription factors, RNA polymerase II transcription initiation and pulmonary dendritic cell development, even though neurological terms (spinocerebellar ataxia) were retained. The x-axis indicates the enrichment significance, while the y-axis described the pathway or term with bubble size indicating the number of genes per pathway.

## Discussion

We found an intriguing pattern of associations of both specific STR loci as well as overall STR burden across the genome with brain structure. Moreover, whereas some STR loci demonstrated global effects on brain volume, others exhibited region-specific effects, indicating a key role for STRs in the modulation of brain structure. Specifically, we observed that individuals with a longer CA motif in an intronic region of the *PRR14L* gene had significantly larger thalamic volume, while repeat number variations of an AATG motif in *NADK* was significantly associated with reduced subcortical gray matter volume. Interestingly, we also found that a higher repeat expansion burden, even just 5 units longer than the human reference, was consistently associated with *larger* volumes across multiple brain regions.

We found that STR variations in the *PRR14L* gene were associated with larger thalamic volume. Even though the function of *PRR14L* in neuronal tissues is yet to be defined, prior studies demonstrated that it is involved in cell division and fate specification (*29*). This raises the possibility that STR variations could influence thalamic development through altered gene regulation or neuronal differentiation. This finding aligns with emerging literature demonstrating the impact of STRs on brain structure and risk for neurological disease (*16, 30*). Given the importance of thalamic volume in sensory processing, cognition, and neuropsychiatric health (*31*), this warrants further investigation into the molecular mechanisms through which *PRR14L* STRs might affect neural development. Moreover, we found that STR polymorphisms in the *NADK* gene were associated with reduced subcortical gray matter volume. In the brain and other tissues, *NADK* is the sole enzyme responsible for phosphorylating NAD+ to produce NADP+. This enzymatic step is critical for maintaining cellular redox homeostasis and supports numerous anabolic and metabolic processes that rely on NADPH-dependent pathways (*32*). This finding suggests that there might be a genetic link between the central NAD+ metabolic pathway and structural brain changes. This link might be particularly relevant as dysregulation of NAD+ and its derivatives has been implicated in aging and neurodegenerative disorders (*33*).

In the last two decades, STRs have increasingly been recognized for their substantial contribution to the genetic basis of many traits across diverse species. Multiple studies have demonstrated that non-pathogenic STR polymorphisms modulate functional traits (*34, 35*). Among the most well-studied STR loci are those located in the period gene of Drosophila melanogaster (thought to participate in circadian rhythmicity), the *Avpr1a* gene of voles (reported to be involved in social recognition memory, aggression and sociosexual behaviours in vertebrates), and the *SLC6A4* gene (implicated in the multiple aspects of primate behaviour) (*36–38*). Moreover, variation in STR lengths within key developmental genes such as *Runx-2* and *Alx-4* have been associated with striking differences in skull shape and limb morphology among dog breeds (*39, 40*). Together, these examples underscore STR variation as a potent mechanism for rapid and flexible phenotypic evolution across species.

Despite advances in animal models, research on the impact of STRs on human brain morphology is scarce, particularly regarding the polygenic burden of STRs across genes with unknown pathogenic thresholds. Our study provides novel evidence that even modest expansions of STRs, within the non-pathogenic range, are associated with increased brain volume (both global and regional) in healthy individuals. Remarkably, we found that increases of even just five STR units in the targeted genomic regions were associated with structural benefits. This association suggests that STRs at sub-pathogenic repeat lengths may promote advantageous developmental changes, potentially by modulating gene expression or protein function during early neurodevelopment (*7*). Indeed, our findings demonstrate that STRs, once referred to as “junk DNA”, are important genetic determinants of brain structure, and thereby may also contribute to human neurodiversity.

An additional intriguing aspect of our results is the clear age-related pattern of STR effects on the brain. In both cohorts, the association between polygenic STR burden (for moderate expansions) and brain volume increased steadily with age, with peak effect size occurring between ages 60 and 70 years. The age-dependent findings suggest that STRs may confer beneficial effects on brain structure during mid-to late adulthood, but that their influence could diminish or even become detrimental at older age. This age-dependent shift aligns with the concept of antagonistic pleiotropy, where genetic variants confer advantages early in life but may lead to adverse outcomes later on (*15, 41*). For STRs, this could mean that moderate expansions support healthy brain development and function. Over time, however, the accumulation of additional repeat units with age, potentially driven by ongoing mutations or somatic instability, may ultimately shift the balance. This concept is supported by findings from the Kids-HD study, which followed children and young adults carrying pathogenic CAG expansions in the *HTT* gene (*13*). Despite their increased risk for HD later in life, these individuals demonstrated significantly better cognitive, behavioral, and motor performance during youth compared to non-carriers. These results support the notion that pathogenic repeat expansions in the *HTT* gene may confer neurodevelopmental advantages early in life, even though these same expansions ultimately lead to deleterious effects in adulthood. It is therefore similarly conceivable that extreme repeat expansions in the genes examined here might also produce detrimental effects with advanced age, albeit at a slower pace compared to alleles in the pathogenic range.

Our enrichment analysis revealed that moderate repeat expansions, common among many participants, were primarily associated with neurological traits as well as several fundamental molecular and cellular processes. This suggests that the spectrum of biological effects of these STRs may depend on expansion size. Notably, we observed enrichment of “genetic anticipation” gene sets among modestly expanded STR loci. This supports the idea that even repeat lengths below the known pathogenic thresholds can engage biological pathways typically linked to repeat expansion disorders (*42*). Intriguingly, individuals with moderately expanded repeats (≥5 units longer than the reference) exhibited increased brain volumes in mid-to late adulthood. Given that this effect diminishes with age, these alleles may exert subtle, age-dependent influences on neurodevelopment and aging, mirroring mechanisms involved in genetic anticipation but without reaching pathological levels.

Among the moderately expanded STRs consistently associated with brain structural measures was one locus within the runt-related transcription factor 2 (*RUNX2*) gene*. RUNX2* encodes a master transcription factor that acts like a “switch” and regulates essential processes such as ossification, cranial suture closure, and bone morphogenesis (*43*). Recent work in dogs revealed that STR variations in *RUNX2* correlate with distinct differences in limb and skull morphology (*44*). Interestingly, *RUNX2* is expressed in key regions of the human brain such as the cerebellum, hippocampus and neocortex (https://www.proteinatlas.org/ENSG00000124813-RUNX2/brain; accessed on July 28, 2025), suggesting that this gene may be implicated in neurogenesis and neuronal function. Our findings further imply a potential role for *RUNX2* in neuronal function via its STR domain, demonstrating that non-pathogenic STR variation is biologically relevant.

Some association signals observed in the Rhineland Study cohort could not be replicated in the UKB. A key factor likely contributing to this discrepancy is the technical difference in sequencing read length between the two datasets. In the Rhineland Study we used a targeted sequencing approach with relatively long paired-end reads (2×250 bp) with a high coverage (∼180) per locus to obtain more accurate estimates of STR sizes, whereas in the UKB cohort WGS with shorter paired-end reads (2×75 bp) and a lower coverage (∼30) was used. The relatively shorter read length and lower coverage per locus in the UKB cohort is likely to have influenced the accuracy of detecting longer repeat expansions, which could partly explain the limited replication of certain associations. In addition, other differences between the two cohorts in demographic composition, especially age and ancestry, may also have contributed to some of the discrepancies. For example, the Rhineland Study sample had a wider age range distribution than the UKB sample, which may have led to underestimation of age-dependent STR effects in the latter. Other limitations of our study include lack of longitudinal and cross-ancestry data, which will be subject of future research.

In conclusion, our work demonstrates that STR polymorphisms can act as important genetic modifiers of brain structure in the general population. Moreover, our findings support the antagonistic pleiotropy model for STRs, indicating that highly polymorphic STR loci may have evolved due to their positive contributions to neuroanatomical plasticity and cognitive function. Future studies exploring the mechanistic basis of these effects are warranted. This especially includes investigating how subtle STR expansions influence gene expression and neurodevelopment. Taken together our work lays the foundation for integrating STR analysis into studies of age-associated brain health and neurodegenerative diseases.

## Materials and Methods

### Experimental Design

This population-based research was conducted using two well-characterized cohorts to investigate the contribution of STR variation to individual differences in brain structure. The study followed a two-stage design: discovery in the Rhineland Study cohort (N=2,958) and replication in the UKB cohort (N=38,879). With this setup, we aimed to identify specific STR loci and polygenic STR patterns associated with brain imaging-derived phenotypes.

### Cohorts Description

#### Rhineland Study

We used data from the first 2,958 participants of the Rhineland Study – an ongoing population-based cohort study in Bonn, Germany – in whom both DNA samples and brain imaging-data were available. The Rhineland Study recruits individuals from two geographically defined districts in Bonn and aims to investigate the etiology, risk factors and predictive biomarkers of aging and age-associated (brain) diseases (*45*). Eligible participants aged 30 years and older are invited to the study center, where they undergo comprehensive physical examinations and in-depth (endo)phenotyping. To ensure clear understanding of the study procedures and informed consent, sufficient proficiency in German is required for inclusion. The protocol of Rhineland Study was approved by the ethics committee of the University of Bonn Medical Faculty (Ref: 338/15). The study is conducted according to the International Conference on Harmonization Good Clinical Practice standards (ICH-GCP), with written informed consent obtained in accordance with the Declaration of Helsinki.

#### UK Biobank Study

The UKB is one of the largest population-based cohort studies to date, including over 500,000 participants. The study was launched in 2006 with invitations sent to 9.2 million eligible individuals aged 40–69 years living in England, Scotland, and Wales. Approximately 500,000 participants enrolled by attending one of 22 assessment centers across the UK, where baseline data were collected through touchscreen questionnaires, verbal interviews, and physical assessments. Biological samples were also obtained to enable genotyping and a wide range of hematological and biochemical assays(*46, 47*). In 2014, the UKB launched its imaging initiative, inviting participants back for multimodal imaging assessments as part of a follow-up visit(*19*). For the present study, we included 49,304 participants who completed their first imaging visit, of whom 45,306 also had WGS data available. We chose the UKB for replication due to its large sample size and the availability of high-quality, harmonized imaging and genomic data. Approval for conducting UKB Study was received from the National Information Governance Board for Health and Social Care and the National Health Service North West Centre for Research Ethics Committee (Ref: 11/NW/0382). Every participant provided written informed consent.

### Targeted STR panel design

For participants of the Rhineland Study with available DNA samples obtained from whole blood, an amplicon-based targeted sequencing approach was applied for the genotyping of approximately 2940 TR-containing regions. These STR regions were curated based on the following criteria: a previously reported causal role in a particular (repeat expansion) disease, potential involvement in brain structural development and function based on comparative genomic studies (*17, 18, 21–24, 48–52*). Additionally, highly polymorphic microsatellite regions that are often utilized in DNA fingerprinting such as the Marshfield panel were also included in the selection panel. Subsequently, a custom targeted amplicon panel targeting these loci was designed in collaboration with Illumina Concierge Design Services using DesignStudio to maximize in-silico coverage and optimize primer configurations. All loci and assay regions were selected to ensure comprehensive coverage of the intended targets.

### DNA extraction and library preparation

Genomic DNA was extracted from peripheral blood samples of 2958 participants of the Rhineland Study using standard magnetic bead-based protocols and quantified through Qubit fluorometry (*53*). Libraries were prepared using the AmpliSeq Library PLUS kit (Illumina, 384 reactions per kit) following the manufacturer’s recommendations (*54*). For each reaction, 50 - 100 ng of genomic DNA was used as input. Multiplex PCR was performed for targeted amplification of the panel regions. Following amplification, PCR products underwent enzymatic primer digestion and magnetic bead-based purification. Purified amplicons were indexed using the AmpliSeq CD Index Set A-D, enabling assignment of unique dual indices to 96 samples per set to facilitate multiplexing. Indexed libraries were quantified (Qubit) and normalized for pooling. Equal molar pools were prepared for sequencing following Illumina’s guidelines. We measured the size-distribution using the Agilent high sensitivity D5000 assay on a TapeStation 4200 system (Agilent technologies).

### Targeted sequencing

Pooled libraries were sequenced on an Illumina NovaSeq 6000 platform using the SP Reagent Kit (500 cycles with 2×250 bp paired-end reads), generating paired-end reads with a targeted average sequencing depth of 180x. All sequencing procedures were performed according to the manufacturer’s workflow for denaturation, dilution, and loading (Illumina, NovaSeq 6000 System Guide). Quality control was performed on the FASTQ files using FastQC with default parameters (Version 0.11.9), after which the high quality reads were aligned to the human reference genome (GRCh38) using the BWA-mem algorithm (version 0.7.17) (*55, 56*). The diploid genotypes in all 2940 loci were then estimated with ExpansionHunter version 5 using our custom variant catalog (*57*).

### UK Biobank whole genome sequencing protocol

WGS of UKB samples was performed using the Illumina NovaSeq platform with mean coverage of 32.5 per sample and 2 × 75 bp paired-end reads. Sequence reads were mapped to the GRCh38 human reference genome using the BWA algorithm. The study design, sample selection, and bioinformatic pipeline have been extensively detailed previously (*20, 58*). To perform STR genotyping in the UKB cohort, we implemented ExpansionHunter as a custom applet and deployed it through the DNAnexus toolkit on the UKB Research Analysis Platform. This setup enabled efficient and scalable genotyping of STRs directly from the aligned WGS data.

### Custom variant catalog design for targeted STR genotyping

The ExpansionHunter algorithm requires a variant catalog, which is a structured JSON file that specifies the genomic coordinates, locus structure, and repeat motif of the loci to be genotyped. Publicly available catalogs exist for genome-wide STR genotyping and for disease-specific pathogenic loci. However, to reduce computational demands and restrict analyses to loci of interest, we generated a custom variant catalog. To build this catalog, we first identified genomic intervals containing exact repeats of DNA motifs (1–10 bp in length) across the autosomes of the GRCh38 reference genome. This was achieved using our in-house tool STRfinder/STRbean (v1.0), applied to the selected STR targets (*28, 59*). To filter out low quality STR loci, we implemented a filtering step in STRfinder using the sequence complexity of flanking regions. Sequence complexity was estimated using Shannon entropy and sequence repetitiveness, and loci with low diversity were excluded from downstream analyses (*60*). Each identified locus was then annotated based on GENCODE v38 (https://www.gencodegenes.org/human/release_38.html). The STRfinder output, generated in BED format, was subsequently converted into a JSON-formatted variant catalog compatible with ExpansionHunter using customized scripts (*61*). The final catalog contained 2,817 polymorphic STR loci, which were then used for downstream genotyping. Notably, this customized catalog was consistent with those produced by recently published STR catalog generation protocols (*61*).

### Quality-control of genotyped STRs

The approach utilized in the STR genotyping and filtering steps followed established methodologies (*62–64*). Briefly, we run ExpansionHunter v5.0.0 separately on each individual BAM file with default parameters using our custom variant catalog (as described above). The individual ExpansionHunter VCF outputs were subsequently sorted, compressed and indexed using bcftools (version 1.15.1) (*65*). The indexed files were further processed using the DumpSTR algorithm from the TRTool Suite (6.1.0) to perform call-level and locus-level filtering as published (*63, 66*). Additionally, the parameter --eh-min-call-LC 10 was included in DumpSTR algorithm to remove low quality calls. Taken together, these steps effectively removed low quality STRs as well as loci whose genotypes did not follow Hardy-Weinberg Equilibrium (at p < 0.001). Moreover, as an additional quality-control step, we applied REViewer (version 0.2.7) to a subset of randomly selected ExpansionHunter output files to visualize read alignments at individual loci (*67*). This step was particularly important for evaluating genotype calls where the repeat region exceeded the read length or where the confidence intervals were wide, enabling a more accurate assessment of genotyping quality. The same STR genotyping and quality-control pipelines were applied to both cohorts.

### Outlier and rare expansion detection

STR genotyping benefits from longer sequencing read lengths to accurately resolve repeat tracts. In the Rhineland Study, sequencing reads were approximately three times longer than those in the UKB. However, challenges in accurately calling repeat lengths persisted particularly for expansions approaching or exceeding the read length. To enhance the reliability of STR calls, we employed a density-based spatial clustering of applications with noise (DBSCAN) approach, as described previously (*26*). DBSCAN is a non-parametric clustering algorithm that identifies dense clusters of data points - here, repeat length estimates - while flagging isolated points in low-density regions as potential outliers. This method enabled robust identification of individuals harboring unusually long STR alleles for each locus, thereby minimizing the inclusion of spurious or artefactual repeat calls.

### Polygenic STR burden

To assess polygenic STR burden, we performed a stratified analysis using predefined tract length thresholds of ≥1, ≥5, ≥10, and ≥20 repeat units longer than the GRCh38 reference genome. We considered the longer of the 2 alleles at each locus. At each threshold, we quantified the number of STR loci exceeding the specified repeat length for each individual which we referred to as polygenic STR expansion burden metric. This score was then tested for association with brain imaging-derived phenotypes while controlling for covariates stated in the statistical analysis section.

### Imaging data

In the Rhineland Study, we used a bespoke imaging protocol to acquire MRI images from participants at two study sites. This involved the use of two identical 3T MRI scanners, each equipped with 64-channel head-neck coils (MAGNETOM Prisma; Siemens Healthcare). The detailed imaging protocol of the Rhineland Study has been described previously(*68*). In this study, we utilized the T1-weighted (T1w), and T2-weighted (T2w) scans.

Participants from the UKB imaging sub-study were invited back to three specialized imaging facilities, each equipped with a 3T Siemens Skyra (software platform VD13) (*69*). Brain imaging was performed using the 3T system with a 32-channel receive head coil (*70*). Detailed imaging protocol of the UKB have been previously published (*71*).

Brain imaging-derived phenotypes were obtained using fully automated MRI segmentation pipelines. In the Rhineland Study, T1w MR images were processed with FreeSurfer, v6.0 (*72*), while in the UKB the FastSurfer pipeline v2.1.0(*73*) was applied to generate the corresponding brain imaging-derived phenotypes.

### Gene ontology enrichment analysis

GO enrichment analysis was conducted in the R statistical environment (v4.1.0). Prior to this, we performed a genome-wide annotation of approximately 1.2 million STRs identified in the GRCh38 human reference genome. To determine the genomic context of each STR, we performed a proximity search against annotated human genes from the UCSC Genome Browser (GRCh38 reference genome) using the TxDb.Hsapiens.UCSC.hg38.knownGene package. The goal was to identify the nearest gene for each STR locus. Only STRs located within a maximum distance of 250 kb from a gene’s TSS were retained for further analysis. This approach ensured that the analysis focused on potentially regulatory or functionally relevant loci. Finally, the Entrez IDs of these nearest genes were subsequently mapped to their official gene symbols using the org.Hs.eg.db annotation package (*74*). This method ensured that all STRs were associated with their closest gene and that the gene names were standardized for downstream interpretation. Next, we evaluated the genes in our custom STR panel at predefined expansion thresholds based on carrier frequency within each cohort. At each threshold (≥1, ≥5, ≥10, or ≥20 repeat units longer than the reference), we identified the most frequent genes shared between cohorts and used these for gene set enrichment analysis. Subsequently, functional enrichment analysis was performed using the selected genes at the respective thresholds as input using gprofiler2 (v0.2.3) (*75*). A custom background gene set, comprising all genes associated with the original STR reference panel, was used to ensure enrichment tests were limited to the relevant genomic space. This over-representation analysis was conducted against multiple databases, including GO Biological Process, GO Molecular Function, GO Cellular Component, the Kyoto Encyclopedia of Genes and Genomes (KEGG) and Reactome. Enrichment of GO and pathway terms was assessed using a cumulative hypergeometric test, and statistical significance was defined as p < 0.05 after correction for multiple testing using the FDR method.

## Statistical analyses

Demographic characteristics were summarized as means with standard deviations, while sex was reported as percentages. For the single association testing we used allele dosages, which was calculated as the mean of the two STR alleles genotyped at each locus. Since most of the STR genotypes were not normally distributed, we first applied rank-based inverse normal transformation to these genotypes. We then performed multiple linear regression to assess the association between STR dosage and brain imaging-derived phenotypes, using STR dosage as a continuous variable (main independent variable) and the brain imaging-derived phenotypes as dependent variables. In each model, we adjusted for age, sex, the first 10 genetic principal components to account for population structure, and only for volumetric measures, additionally for extracted total intracranial volume to account for differences in head size. We also included a quadratic term for age to account for potential non-linear relationships between the STRs and the brain imaging-derived phenotypes. We report the estimates and p-values of the rank-based inverse normal transformed STR genotypes. To correct for multiple comparisons across the tested loci, we applied Bonferroni correction by setting the significance threshold to p < 0.05/2477 for the number of independent loci tested. For the polygenic STR burden, because each threshold was evaluated independently, we applied the Li & Ji method to correct for multiple testing. This method accounts for the correlation among the brain imaging-derived phenotypes. Based on the number of effective tests, the statistical significance threshold was set at 0.05/26 for the Rhineland Study and 0.05/22 for the UKB. Similarly, the threshold for replication analyses was determined according to the number of top hits selected from the discovery cohort. The significance threshold for the replication analyses depended on the number of loci selected for replication. The significance threshold was p<0.05/n, where n is the number of hits selected for replication.

## Data Availability

Due to data protection regulations, the Rhineland Study data are not publicly available. However, qualified researchers may request access in accordance with the Rhineland Study Data Use and Access Policy. Requests for data access or additional information can be directed to. All whole-genome sequencing as well as individual-level data from the UK Biobank Imaging Study used in this work are available through the UKB Resource (https://www.ukbiobank.ac.uk). Access to these data is subject to UK Biobank data access policies. Part of this research has been conducted using the UK Biobank Resource under Application Number 82056. All data are available in the main text or the supplementary materials.

## Acknowledgments

We would like to thank the Rhineland Study team for supporting the data acquisition and management as well as the PRECISE team for sequencing support.

## Author contributions

Conceptualization: NAA, MMBB, RM

Methodology: RM, NAA, MMBB, JH, MDB, EDD, RS, KH, SE, MT, TB, AMS

Data analysis: RM, JH, SE, NAA Supervision: NAA, MMBB Writing–original draft: RM, NAA

Writing–review & editing: RM, NAA, MMBB, RM, JH, MDB, EDD, RS, KH, SE, MT, TB, AMS

Funding acquisition – NAA, MMBB

## Data and materials availability

Due to data protection regulations, the Rhineland Study data are not publicly available. However, qualified researchers may request access in accordance with the Rhineland Study’s Data Use and Access Policy. Requests for data access or additional information can be directed to. All whole-genome sequencing as well as individual-level data from the UK Biobank Imaging Study used in this work are available through the UKB Resource (https://www.ukbiobank.ac.uk). Access to these data is subject to UK Biobank’s data access policies. Part of this research has been conducted using the UK Biobank Resource under Application Number 82056. All data are available in the main text or the supplementary materials.

## References

1. A. Abdellaoui, L. Yengo, K. J. H. Verweij, P. M. Visscher, 15 years of GWAS discovery: Realizing the promise. Am J Hum Genet 110, 179–194 (2023).

2. J. Hardy, Escott-Price, V, The genetics of neurodegenerative diseases is the genetics of age-related damage clearance failure. Mol Psychiatry 30, 2748–2753 (2025).

3. J. C. Lambert, C. A. Ibrahim-Verbaas, D. Harold, A. C. Naj, R. Sims, C. Bellenguez, A. L. DeStafano, J. C. Bis, G. W. Beecham, B. Grenier-Boley, G. Russo, T. A. Thorton-Wells, N. Jones, A. V. Smith, V. Chouraki, C. Thomas, M. A. Ikram, D. Zelenika, B. N. Vardarajan, Y. Kamatani, C. F. Lin, A. Gerrish, H. Schmidt, B. Kunkle, M. L. Dunstan, A. Ruiz, M. T. Bihoreau, S. H. Choi, C. Reitz, F. Pasquier, C. Cruchaga, D. Craig, N. Amin, C. Berr, O. L. Lopez, P. L. De Jager, V. Deramecourt, J. A. Johnston, D. Evans, S. Lovestone, L. Letenneur, F. J. Moron, D. C. Rubinsztein, G. Eiriksdottir, K. Sleegers, A. M. Goate, N. Fievet, M. W. Huentelman, M. Gill, K. Brown, M. I. Kamboh, L. Keller, P. Barberger-Gateau, B. McGuiness, E. B. Larson, R. Green, A. J. Myers, C. Dufouil, S. Todd, D. Wallon, S. Love, E. Rogaeva, J. Gallacher, P. St George-Hyslop, J. Clarimon, A. Lleo, A. Bayer, D. W. Tsuang, L. Yu, M. Tsolaki, P. Bossu, G. Spalletta, P. Proitsi, J. Collinge, S. Sorbi, F. Sanchez-Garcia, N. C. Fox, J. Hardy, M. C. Deniz Naranjo, P. Bosco, R. Clarke, C. Brayne, D. Galimberti, M. Mancuso, F. Matthews, I. European Alzheimer’s Disease, Genetic, D. Environmental Risk in Alzheimer’s, C. Alzheimer’s Disease Genetic, H. Cohorts for, E. Aging Research in Genomic, S. Moebus, P. Mecocci, M. Del Zompo, W. Maier, H. Hampel, A. Pilotto, M. Bullido, F. Panza, P. Caffarra, B. Nacmias, J. R. Gilbert, M. Mayhaus, L. Lannefelt, H. Hakonarson, S. Pichler, M. M. Carrasquillo, M. Ingelsson, D. Beekly, V. Alvarez, F. Zou, O. Valladares, S. G. Younkin, E. Coto, K. L. Hamilton-Nelson, W. Gu, C. Razquin, P. Pastor, I. Mateo, M. J. Owen, K. M. Faber, P. V. Jonsson, O. Combarros, M. C. O’Donovan, L. B. Cantwell, H. Soininen, D. Blacker, S. Mead, T. H. Mosley, Jr., D. A. Bennett, T. B. Harris, L. Fratiglioni, C. Holmes, R. F. de Bruijn, P. Passmore, T. J. Montine, K. Bettens, J. I. Rotter, A. Brice, K. Morgan, T. M. Foroud, W. A. Kukull, D. Hannequin, J. F. Powell, M. A. Nalls, K. Ritchie, K. L. Lunetta, J. S. Kauwe, E. Boerwinkle, M. Riemenschneider, M. Boada, M. Hiltuenen, E. R. Martin, R. Schmidt, D. Rujescu, L. S. Wang, J. F. Dartigues, R. Mayeux, C. Tzourio, A. Hofman, M. M. Nothen, C. Graff, B. M. Psaty, L. Jones, J. L. Haines, P. A. Holmans, M. Lathrop, M. A. Pericak-Vance, L. J. Launer, L. A. Farrer, C. M. van Duijn, C. Van Broeckhoven, V. Moskvina, S. Seshadri, J. Williams, G. D. Schellenberg, P. Amouyel, Meta-analysis of 74,046 individuals identifies 11 new susceptibility loci for Alzheimer’s disease. Nat Genet 45, 1452–1458 (2013).

4. S. Gandhi, N. W. Wood, Genome-wide association studies: the key to unlocking neurodegeneration? Nat Neurosci 13, 789–794 (2010).

5. H. Fan, J. Y. Chu, A brief review of short tandem repeat mutation. Genomics Proteomics Bioinformatics 5, 7–14 (2007).

6. A. T. M. Bagshaw, Functional Mechanisms of Microsatellite DNA in Eukaryotic Genomes. Genome Biol Evol 9, 2428–2443 (2017).

7. S. F. Fotsing, J. Margoliash, C. Wang, S. Saini, R. Yanicky, S. Shleizer-Burko, A. Goren, M. Gymrek, The impact of short tandem repeat variation on gene expression. Nat Genet 51, 1652–1659 (2019).

8. A. N. Khristich, S. M. Mirkin, On the wrong DNA track: Molecular mechanisms of repeat-mediated genome instability. J Biol Chem 295, 4134–4170 (2020).

9. M. O. Press, K. D. Carlson, C. Queitsch, The overdue promise of short tandem repeat variation for heritability. Trends Genet 30, 504–512 (2014).

10. S. L. Gardiner, S. Trompet, B. Sabayan, M. W. Boogaard, J. W. Jukema, P. E. Slagboom, R. A. C. Roos, J. van der Grond, N. A. Aziz, Repeat variations in polyglutamine disease-associated genes and cognitive function in old age. Neurobiol Aging 84, 236 e217–236 e228 (2019).

11. S. L. Gardiner, M. J. van Belzen, M. W. Boogaard, W. M. C. van Roon-Mom, M. P. Rozing, A. M. van Hemert, J. H. Smit, A. T. F. Beekman, G. van Grootheest, R. A. Schoevers, R. C. Oude Voshaar, H. C. Comijs, B. Penninx, R. C. van der Mast, R. A. C. Roos, N. A. Aziz, Large normal-range TBP and ATXN7 CAG repeat lengths are associated with increased lifetime risk of depression. Transl Psychiatry 7, e1143 (2017).

12. S. L. Gardiner, A. V. E. Harder, Y. J. M. Campman, S. Trompet, J. Gussekloo, M. J. van Belzen, M. W. Boogaard, R. A. C. Roos, I. E. Jansen, Y. A. L. Pijnenburg, P. Scheltens, W. M. van der Flier, N. A. Aziz, Repeat length variations in ATXN1 and AR modify disease expression in Alzheimer’s disease. Neurobiol Aging 73, 230 e239–230 e217 (2019).

13. M. Neema, J. L. Schultz, D. R. Langbehn, A. L. Conrad, E. A. Epping, V. A. Magnotta, P. C. Nopoulos, Mutant Huntingtin Drives Development of an Advantageous Brain Early in Life: Evidence in Support of Antagonistic Pleiotropy. Ann Neurol 96, 1006–1019 (2024).

14. W. Qian, D. Ma, C. Xiao, Z. Wang, J. Zhang, The genomic landscape and evolutionary resolution of antagonistic pleiotropy in yeast. Cell Rep 2, 1399–1410 (2012).

15. G. C. Williams, Pleiotropy, natural selection, and the evolution of senescence. Evolution 11, 398–411 (1957).

16. Q. Liu, W. Tian, Association of human-specific expanded short tandem repeats with neuron-specific regulatory features. Sci Adv 11, eadp9707 (2025).

17. E. J. Vallender, N. Mekel-Bobrov, B. T. Lahn, Genetic basis of human brain evolution. Trends Neurosci 31, 637–644 (2008).

18. T. Bilgin Sonay, T. Carvalho, M. D. Robinson, M. P. Greminger, M. Krutzen, D. Comas, G. Highnam, D. Mittelman, A. Sharp, T. Marques-Bonet, A. Wagner, Tandem repeat variation in human and great ape populations and its impact on gene expression divergence. Genome Res 25, 1591–1599 (2015).

19. T. J. Littlejohns, J. Holliday, L. M. Gibson, S. Garratt, N. Oesingmann, F. Alfaro-Almagro, J. D. Bell, C. Boultwood, R. Collins, M. C. Conroy, N. Crabtree, N. Doherty, A. F. Frangi, N. C. Harvey, P. Leeson, K. L. Miller, S. Neubauer, S. E. Petersen, J. Sellors, S. Sheard, S. M. Smith, C. L. M. Sudlow, P. M. Matthews, N. E. Allen, The UK Biobank imaging enhancement of 100,000 participants: rationale, data collection, management and future directions. Nat Commun 11, 2624 (2020).

20. U. K. B. W.-G. S. Consortium, Whole-genome sequencing of 490,640 UK Biobank participants. Nature, (2025).

21. M. M. Course, K. Gudsnuk, S. N. Smukowski, K. Winston, N. Desai, J. P. Ross, A. Sulovari, C. V. Bourassa, D. Spiegelman, J. Couthouis, C. E. Yu, D. W. Tsuang, S. Jayadev, M. A. Kay, A. D. Gitler, N. Dupre, E. E. Eichler, P. A. Dion, G. A. Rouleau, P. N. Valdmanis, Evolution of a Human-Specific Tandem Repeat Associated with ALS. Am J Hum Genet 107, 445–460 (2020).

22. E. J. Vowles, W. Amos, Evidence for widespread convergent evolution around human microsatellites. PLoS Biol 2, E199 (2004).

23. G. Roth, U. Dicke, Evolution of the brain and intelligence. Trends Cogn Sci 9, 250–257 (2005).

24. K. Kim, S. Bang, D. Yoo, H. Kim, S. Suzuki, De novo emergence and potential function of human-specific tandem repeats in brain-related loci. Hum Genet 138, 661–672 (2019).

25. K. W. Broman, J. C. Murray, V. C. Sheffield, R. L. White, J. L. Weber, Comprehensive human genetic maps: individual and sex-specific variation in recombination. Am J Hum Genet 63, 861–869 (1998).

26. M. H. Guo, W. P. Lee, B. Vardarajan, G. D. Schellenberg, J. E. Phillips-Cremins, Polygenic burden of short tandem repeat expansions promotes risk for Alzheimer’s disease. Nat Commun 16, 1126 (2025).

27. J. Li, L. Ji, Adjusting multiple testing in multilocus analyses using the eigenvalues of a correlation matrix. Heredity (Edinb) 95, 221–227 (2005).

28. J. Hu, A. Aziz, A comprehensive catalog of exact short tandem repeat regions on autosomes and sex chromosomes of the human genome GRCh38. Zenodo, (2024).

29. A. Chase, A. Pellagatti, S. Singh, J. Score, W. J. Tapper, F. Lin, Y. Hoade, C. Bryant, N. Trim, B. H. Yip, K. Zoi, C. Rasi, L. A. Forsberg, J. P. Dumanski, J. Boultwood, N. C. P. Cross, PRR14L mutations are associated with chromosome 22 acquired uniparental disomy, age-related clonal hematopoiesis and myeloid neoplasia. Leukemia 33, 1184–1194 (2019).

30. S. E. Wright, P. K. Todd, Native functions of short tandem repeats. Elife 12, (2023).

31. M. Alhesain, A. Alzu’bi, N. Sankar, C. Smith, J. Kerwin, R. Laws, S. Lindsay, G. J. Clowry, Development of the early fetal human thalamus: from a protomap to emergent thalamic nuclei. Front Neuroanat 19, 1530236 (2025).

32. K. Ohashi, S. Kawai, K. Murata, Identification and characterization of a human mitochondrial NAD kinase. Nat Commun 3, 1248 (2012).

33. N. Xie, L. Zhang, W. Gao, C. Huang, P. E. Huber, X. Zhou, C. Li, G. Shen, B. Zou, NAD(+) metabolism: pathophysiologic mechanisms and therapeutic potential. Signal Transduct Target Ther 5, 227 (2020).

34. M. Verbiest, M. Maksimov, Y. Jin, M. Anisimova, M. Gymrek, T. Bilgin Sonay, Mutation and selection processes regulating short tandem repeats give rise to genetic and phenotypic diversity across species. J Evol Biol 36, 321–336 (2023).

35. M. Gymrek, T. Willems, A. Guilmatre, H. Zeng, B. Markus, S. Georgiev, M. J. Daly, A. L. Price, J. K. Pritchard, A. J. Sharp, Y. Erlich, Abundant contribution of short tandem repeats to gene expression variation in humans. Nat Genet 48, 22–29 (2016).

36. J. M. Ding, Molecular Genetics of Circadian Rhythms in Drosophila. Annu Rev Neurosci, (1998).

37. K. P. Lesch, D. Bengel, A. Heils, S. Z. Sabol, B. D. Greenberg, S. Petri, J. Benjamin, C. R. Muller, D. H. Hamer, D. L. Murphy, Association of anxiety-related traits with a polymorphism in the serotonin transporter gene regulatory region. Science 274, 1527–1531 (1996).

38. E. A. Hammock, L. J. Young, Microsatellite instability generates diversity in brain and sociobehavioral traits. Science 308, 1630–1634 (2005).

39. T. B. Ritzman, N. Banovich, K. P. Buss, J. Guida, M. A. Rubel, J. Pinney, B. Khang, M. J. Ravosa, A. C. Stone, Facing the facts: The Runx2 gene is associated with variation in facial morphology in primates. J Hum Evol 111, 139–151 (2017).

40. J. G. Fondon, Harold, Molecular origins of rapid and continuous morphological evolution. Proceedings of the National Academy of Sciences of the United States of America 101, 18058–18063 (2005).

41. C. Lockwood, A. S. Vo, H. Bellafard, A. J. R. Carter, More evidence for widespread antagonistic pleiotropy in polymorphic disease alleles. Front Genet 15, 1404516 (2024).

42. C. T. McMurray, Mechanisms of trinucleotide repeat instability during human development. Nat Rev Genet 11, 786–799 (2010).

43. T. M. Schroeder, E. D. Jensen, J. J. Westendorf, Runx2: a master organizer of gene transcription in developing and maturing osteoblasts. Birth Defects Res C Embryo Today 75, 213–225 (2005).

44. M. A. Pointer, J. M. Kamilar, V. Warmuth, S. G. Chester, F. Delsuc, N. I. Mundy, R. J. Asher, B. J. Bradley, RUNX2 tandem repeats and the evolution of facial length in placental mammals. BMC Evol Biol 12, 103 (2012).

45. M. M. B. Breteler, A Stöcker, Tony, A Pracht, Eberhard, A Brenner, Daniel, A Stirnberg, Rüdiger, MRI in the Rhineland study: a novel protocol for population neuroimaging. Alzheimer’s and dementia 10, (2014).

46. A. Fry, T. J. Littlejohns, C. Sudlow, N. Doherty, L. Adamska, T. Sprosen, R. Collins, N. E. Allen, Comparison of Sociodemographic and Health-Related Characteristics of UK Biobank Participants With Those of the General Population. Am J Epidemiol 186, 1026–1034 (2017).

47. C. Bycroft, C. Freeman, D. Petkova, G. Band, L. T. Elliott, K. Sharp, A. Motyer, D. Vukcevic, O. Delaneau, J. O’Connell, A. Cortes, S. Welsh, A. Young, M. Effingham, G. McVean, S. Leslie, N. Allen, P. Donnelly, J. Marchini, The UK Biobank resource with deep phenotyping and genomic data. Nature 562, 203–209 (2018).

48. A. Kirby, A. Gnirke, D. B. Jaffe, V. Baresova, N. Pochet, B. Blumenstiel, C. Ye, D. Aird, C. Stevens, J. T. Robinson, M. N. Cabili, I. Gat-Viks, E. Kelliher, R. Daza, M. DeFelice, H. Hulkova, J. Sovova, P. Vylet’al, C. Antignac, M. Guttman, R. E. Handsaker, D. Perrin, S. Steelman, S. Sigurdsson, S. J. Scheinman, C. Sougnez, K. Cibulskis, M. Parkin, T. Green, E. Rossin, M. C. Zody, R. J. Xavier, M. R. Pollak, S. L. Alper, K. Lindblad-Toh, S. Gabriel, P. S. Hart, A. Regev, C. Nusbaum, S. Kmoch, A. J. Bleyer, E. S. Lander, M. J. Daly, Mutations causing medullary cystic kidney disease type 1 lie in a large VNTR in MUC1 missed by massively parallel sequencing. Nat Genet 45, 299–303 (2013).

49. M. Hijikata, I. Matsushita, G. Tanaka, T. Tsuchiya, H. Ito, K. Tokunaga, J. Ohashi, S. Homma, Y. Kobashi, Y. Taguchi, A. Azuma, S. Kudoh, N. Keicho, Molecular cloning of two novel mucin-like genes in the disease-susceptibility locus for diffuse panbronchiolitis. Hum Genet 129, 117–128 (2011).

50. A. L. Byrd, S. B. Manuck, MAOA, childhood maltreatment, and antisocial behavior: meta-analysis of a gene-environment interaction. Biol Psychiatry 75, 9–17 (2014).

51. S. P. A. Benedetti, Casamassima F, Lattanzi L, Liberti M, Musetti L, Cassano GB, Bipolar disorder in late life: clinical characteristics in a sample of older adults admitted for manic episode. Clin Pract Epidemiol Ment Health 4:22., (2008).

52. A. I. Seixas, J. R. Loureiro, C. Costa, A. Ordonez-Ugalde, H. Marcelino, C. L. Oliveira, J. L. Loureiro, A. Dhingra, E. Brandao, V. T. Cruz, A. Timoteo, B. Quintans, G. A. Rouleau, P. Rizzu, A. Carracedo, J. Bessa, P. Heutink, J. Sequeiros, M. J. Sobrido, P. Coutinho, I. Silveira, A Pentanucleotide ATTTC Repeat Insertion in the Non-coding Region of DAB1, Mapping to SCA37, Causes Spinocerebellar Ataxia. Am J Hum Genet 101, 87–103 (2017).

53. A. Thermo Fisher Scientific, Qubit Fluorometric Quantitation. (2022).

54. A. Illumina Inc, AmpliSeq Library PLUS for Illumina: Reference Guide. (2022).

55. S. Andrews, FastQC: A quality control tool for high throughput sequence data. (2010).

56. H. Li, Aligning sequence reads, clone sequences and assembly contigs with BWA-MEM. arXiv:1303.3997 [q-bio.GN]. (2013).

57. E. Dolzhenko, V. Deshpande, F. Schlesinger, P. Krusche, R. Petrovski, S. Chen, D. Emig-Agius, A. Gross, G. Narzisi, B. Bowman, K. Scheffler, J. van Vugt, C. French, A. Sanchis-Juan, K. Ibanez, A. Tucci, B. R. Lajoie, J. H. Veldink, F. L. Raymond, R. J. Taft, D. R. Bentley, M. A. Eberle, ExpansionHunter: a sequence-graph-based tool to analyze variation in short tandem repeat regions. Bioinformatics 35, 4754–4756 (2019).

58. B. V. Halldorsson, H. P. Eggertsson, K. H. S. Moore, H. Hauswedell, O. Eiriksson, M. O. Ulfarsson, G. Palsson, M. T. Hardarson, A. Oddsson, B. O. Jensson, S. Kristmundsdottir, B. D. Sigurpalsdottir, O. A. Stefansson, D. Beyter, G. Holley, V. Tragante, A. Gylfason, P. I. Olason, F. Zink, M. Asgeirsdottir, S. T. Sverrisson, B. Sigurdsson, S. A. Gudjonsson, G. T. Sigurdsson, G. H. Halldorsson, G. Sveinbjornsson, K. Norland, U. Styrkarsdottir, D. N. Magnusdottir, S. Snorradottir, K. Kristinsson, E. Sobech, H. Jonsson, A. J. Geirsson, I. Olafsson, P. Jonsson, O. B. Pedersen, C. Erikstrup, S. Brunak, S. R. Ostrowski, D. G. Consortium, G. Thorleifsson, F. Jonsson, P. Melsted, I. Jonsdottir, T. Rafnar, H. Holm, H. Stefansson, J. Saemundsdottir, D. F. Gudbjartsson, O. T. Magnusson, G. Masson, U. Thorsteinsdottir, A. Helgason, H. Jonsson, P. Sulem, K. Stefansson, The sequences of 150,119 genomes in the UK Biobank. Nature 607, 732–740 (2022).

59. J. Hu, A. Aziz, A comprehensive catalog of approxiamte short tandem repeat regions on autosomes and sex chromosomes of the human genome GRCh38. (2024).

60. J. Hu, searchSTR and STRfinder in a package STRbean. (2025).

61. A. Halman, A. Lonsdale, A. Oshlack, Analysis of Tandem Repeats in Short-Read Sequencing Data: From Genotyping Known Pathogenic Repeats to Discovering Novel Expansions. Curr Protoc 4, e70010 (2024).

62. M. Gymrek, D. Golan, S. Rosset, Y. Erlich, lobSTR: A short tandem repeat profiler for personal genomes. Genome Res 22, 1154–1162 (2012).

63. N. Mousavi, S. Shleizer-Burko, R. Yanicky, M. Gymrek, Profiling the genome-wide landscape of tandem repeat expansions. Nucleic Acids Res 47, e90 (2019).

64. E. Dolzhenko, J. van Vugt, R. J. Shaw, M. A. Bekritsky, M. van Blitterswijk, G. Narzisi, S. S. Ajay, V. Rajan, B. R. Lajoie, N. H. Johnson, Z. Kingsbury, S. J. Humphray, R. D. Schellevis, W. J. Brands, M. Baker, R. Rademakers, M. Kooyman, G. H. P. Tazelaar, M. A. van Es, R. McLaughlin, W. Sproviero, A. Shatunov, A. Jones, A. Al Khleifat, A. Pittman, S. Morgan, O. Hardiman, A. Al-Chalabi, C. Shaw, B. Smith, E. J. Neo, K. Morrison, P. J. Shaw, C. Reeves, L. Winterkorn, N. S. Wexler, U. S.-V. C. R. Group, D. E. Housman, C. W. Ng, A. L. Li, R. J. Taft, L. H. van den Berg, D. R. Bentley, J. H. Veldink, M. A. Eberle, Detection of long repeat expansions from PCR-free whole-genome sequence data. Genome Res 27, 1895–1903 (2017).

65. P. Danecek, J. K. Bonfield, J. Liddle, J. Marshall, V. Ohan, M. O. Pollard, A. Whitwham, T. Keane, S. A. McCarthy, R. M. Davies, H. Li, Twelve years of SAMtools and BCFtools. Gigascience 10, (2021).

66. Y. Cui, W. Ye, J. S. Li, J. J. Li, E. Vilain, T. Sallam, W. Li, A genome-wide spectrum of tandem repeat expansions in 338,963 humans. Cell 187, 6411–6412 (2024).

67. E. Dolzhenko, B. Weisburd, K. Ibanez, I. S. Rajan-Babu, C. Anyansi, M. F. Bennett, K. Billingsley, A. Carroll, S. Clamons, M. C. Danzi, V. Deshpande, J. Ding, S. Fazal, A. Halman, B. Jadhav, Y. Qiu, P. A. Richmond, C. T. Saunders, K. Scheffler, J. van Vugt, R. Zwamborn, C. Genomics England Research, S. S. Chong, J. M. Friedman, A. Tucci, H. L. Rehm, M. A. Eberle, REViewer: haplotype-resolved visualization of read alignments in and around tandem repeats. Genome Med 14, 84 (2022).

68. A. Koch, R. Stirnberg, S. Estrada, W. Zeng, V. Lohner, M. Shahid, P. Ehses, E. D. Pracht, M. Reuter, T. Stocker, M. M. B. Breteler, Versatile MRI acquisition and processing protocol for population-based neuroimaging. Nat Protoc 20, 1223–1245 (2025).

69. K. L. Miller, F. Alfaro-Almagro, N. K. Bangerter, D. L. Thomas, E. Yacoub, J. Xu, A. J. Bartsch, S. Jbabdi, S. N. Sotiropoulos, J. L. Andersson, L. Griffanti, G. Douaud, T. W. Okell, P. Weale, I. Dragonu, S. Garratt, S. Hudson, R. Collins, M. Jenkinson, P. M. Matthews, S. M. Smith, Multimodal population brain imaging in the UK Biobank prospective epidemiological study. Nat Neurosci 19, 1523–1536 (2016).

70. S. M. Smith, Alfaro-Almagro, F. & Miller, K. L., UK Biobank Brain Imaging Documentation. Oxford Centre for Functional MRI of the Brain (FMRIB/WIN), University of Oxford, (2024).

71. F. Alfaro-Almagro, M. Jenkinson, N. K. Bangerter, J. L. R. Andersson, L. Griffanti, G. Douaud, S. N. Sotiropoulos, S. Jbabdi, M. Hernandez-Fernandez, E. Vallee, D. Vidaurre, M. Webster, P. McCarthy, C. Rorden, A. Daducci, D. C. Alexander, H. Zhang, I. Dragonu, P. M. Matthews, K. L. Miller, S. M. Smith, Image processing and Quality Control for the first 10,000 brain imaging datasets from UK Biobank. Neuroimage 166, 400–424 (2018).

72. B. Fischl, FreeSurfer. Neuroimage 62, 774–781 (2012).

73. L. Henschel, S. Conjeti, S. Estrada, K. Diers, B. Fischl, M. Reuter, FastSurfer - A fast and accurate deep learning based neuroimaging pipeline. Neuroimage 219, 117012 (2020).

74. M. Carlson, org.Hs.eg.db: Genome wide annotation for Human. R package version 3.17.0. Bioconductor, (2023).

75. U. Raudvere, L. Kolberg, I. Kuzmin, T. Arak, P. Adler, H. Peterson, J. Vilo, g:Profiler: a web server for functional enrichment analysis and conversions of gene lists (2019 update). Nucleic Acids Res 47, W191–W198 (2019).

