## Supplementary Material for "Tandem Repeat Polymorphisms Are Associated with Brain Structure: Results of Two Large Population-based Studies"

Richard Mantey *et al.*

**This PDF file includes:**

Supplementary Text

Figs. S1 to S9

Tables S1 to S3

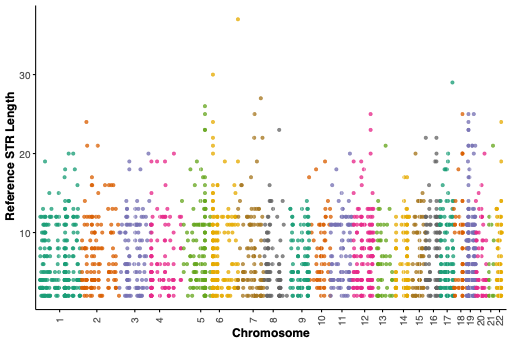

**Fig. S1.** **Distribution of allele lengths for our customized loci in the human reference genome (GRCh38).** The Manhattan plot illustrates the distribution of STR tract lengths across genomic regions represented in our customized panel, as annotated in the human reference genome. Motif lengths ranged from 1 to 10 base pairs, with dinucleotide repeats being the most common, accounting for 36% of all STRs. Mononucleotide repeats constituted 19%, while tri- to hexanucleotide motifs collectively represented approximately 35% of the panel. STRs with motif lengths between 7 and 10 base pairs comprised less than 10% of the panel.

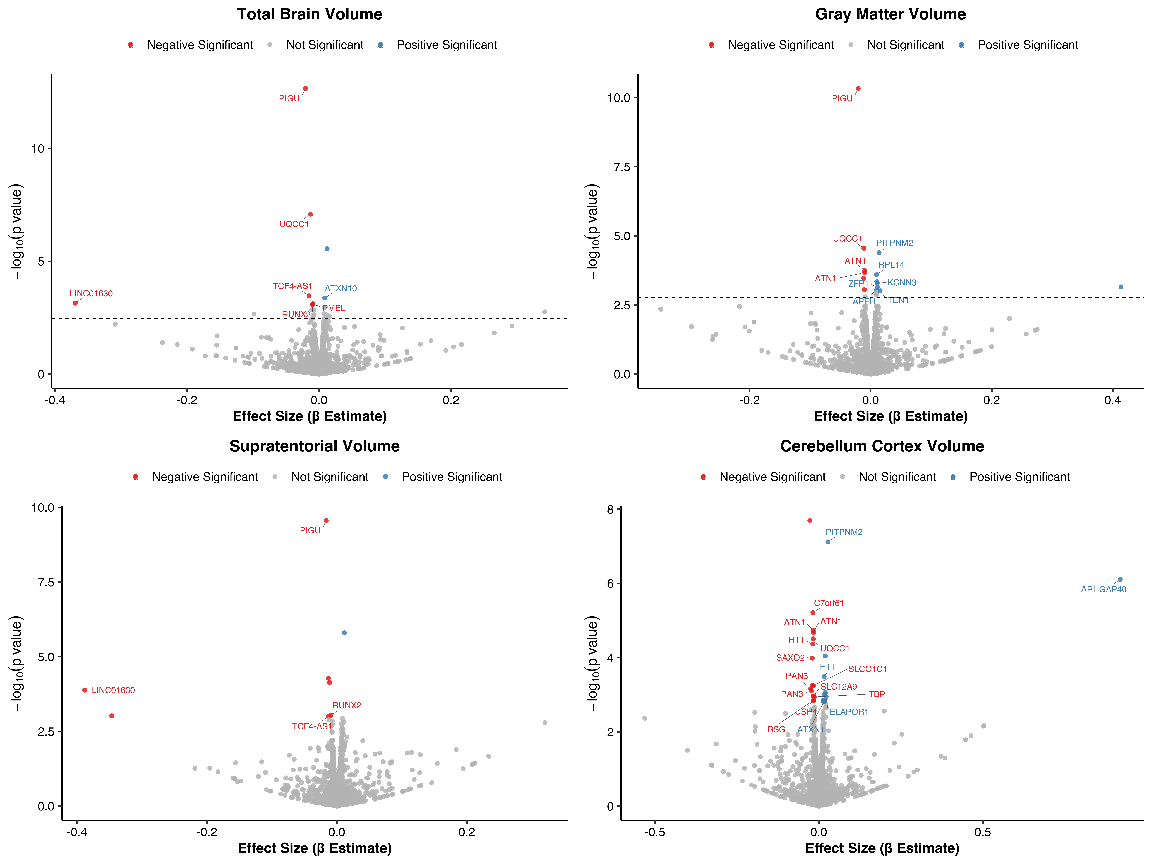

**Fig. S2. Replication results across other brain phenotypes.** This figure shows the replication analysis for STR associations with additional brain phenotypes in the UK Biobank. For total brain volume, 8 of 15 loci showed consistent directions of effect, with several being nominally significant. For total gray matter volume, 20 of 30 loci were directionally consistent, with 3 reaching nominal significance. For supratentorial volume, 3 of 7 loci showed consistent effect direction and magnitude. For cerebellar cortex volume, 1 of 2 loci replicated at nominal significance.

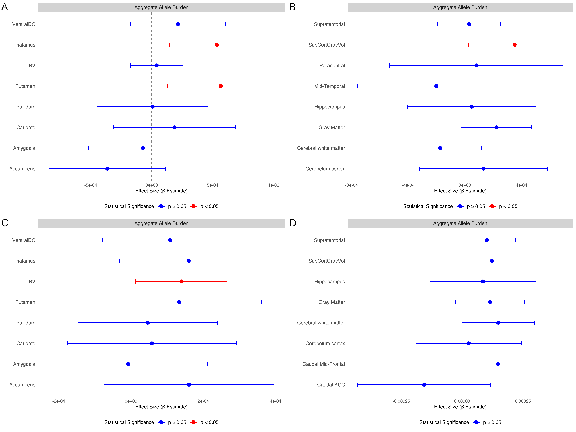

**Fig. S3. Association between aggregate STR allele burden and brain imaging phenotypes across cohorts.** The upper panel shows results for the RS, and the lower panel for the UKB. Significant associations were observed for putamen, thalamic volume, and subcortical gray matter volume. For most brain regions, the direction of association was consistent across cohorts. Overall, higher aggregate allele burden was associated with increased brain volume in multiple regions.

***Abbreviations:*** *eTIV, estimated total intracranial volume; Rostral ACC, rostral anterior cingulate cortex; VentralDC, ventral diencephalon; RS, Rhineland Study; SubCortGrayVol, subcortical gray matter volume; TBV, total brain volume; UKB, UK Biobank.*

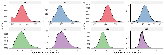

**Fig. S4. Distribution of polygenic STR expansion burden across two cohorts.**

Density plots depict the burden of STR expansions per individual at four thresholds: ≥1, ≥5, ≥10, and ≥20 repeat units longer than the GRCh38 reference genome. The polygenic STR burden for each individual represents the number of loci exceeding the specified threshold. Panels show Rhineland Study (A) and UKB (B). In both cohorts, higher expansion thresholds correspond to fewer loci meeting the criterion, reflecting the rarity of longer STR expansions in the general population. These distributions were used to assess the contribution of STR expansion burden to brain structural variation.

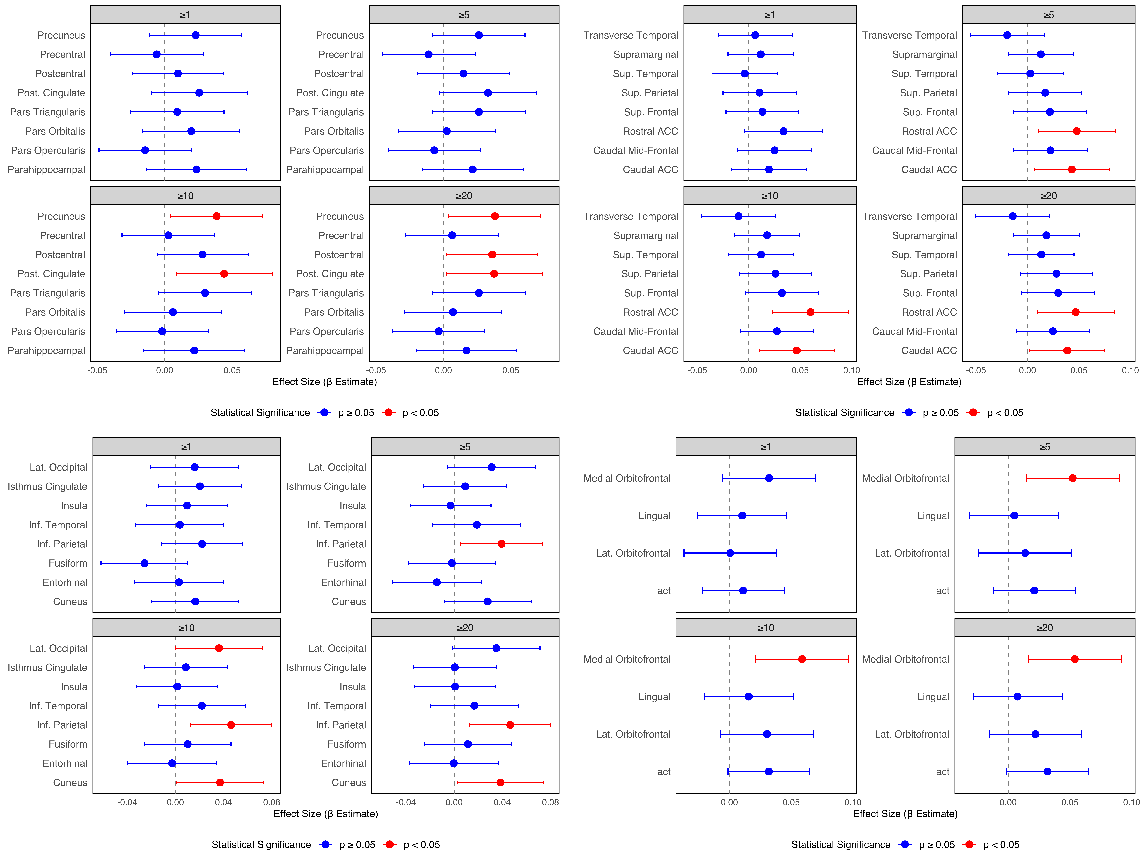

**Fig. S5.** **Forest plot of standardized β estimates (95% CI) for associations between polygenic STR expansion burden and brain phenotypes in the RS discovery cohort.** Each point corresponds to the β estimate for a given brain region at a specific STR threshold, with horizontal bars indicating the 95% confidence interval. Associations significant after Li & Ji multiple-testing correction are shown in red; non-significant associations are shown in blue. The vertical dashed line represents the null effect (β = 0).

***Abbreviations:*** *act, cortical thickness; eTIV, estimated total intracranial volume; Rostral ACC, rostral anterior cingulate cortex; VentralDC, ventral diencephalon*

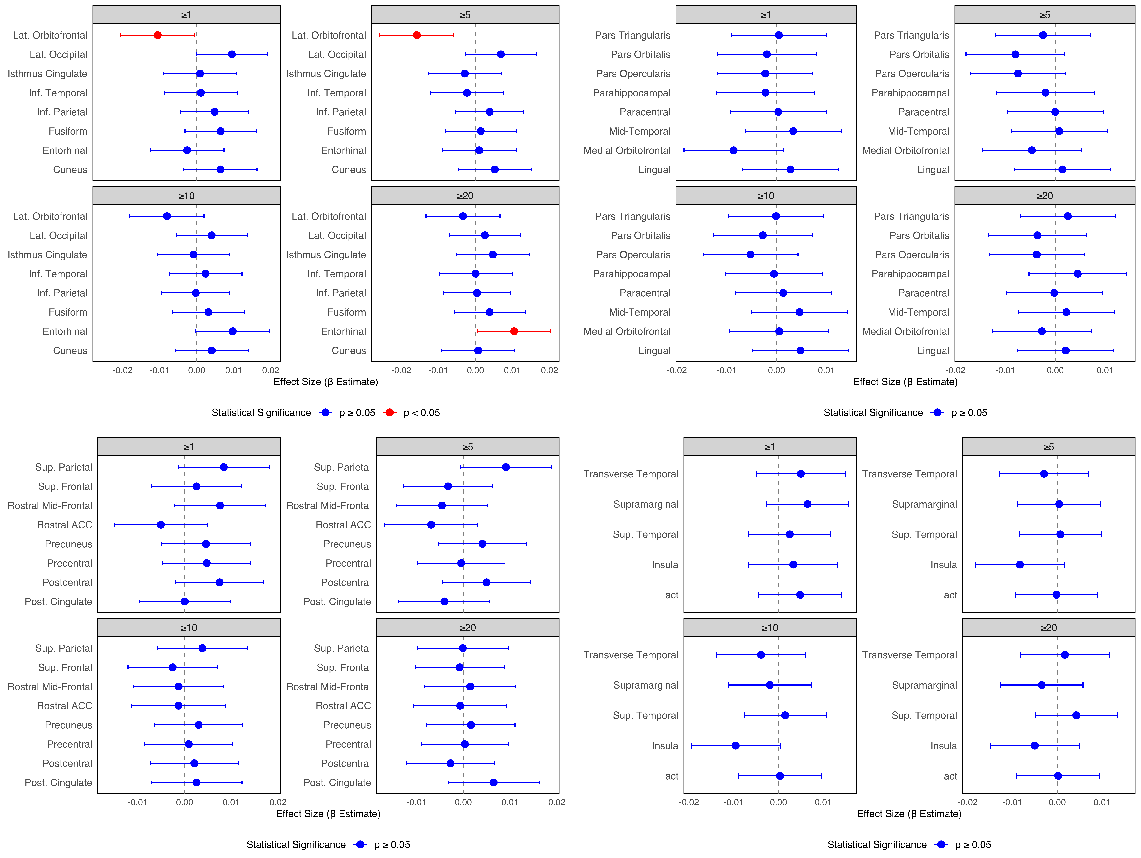

**Fig. S6. Forest plot of standardized β with 95% confidence intervals for the association between polygenic STR expansion burden and brain phenotypes in the UKB replication cohort.** Each point corresponds to the β estimate for a given brain region at a specific STR threshold, with horizontal bars indicating the 95% confidence interval. Associations that survived multiple-testing correction using the Li & Ji are shown in red whereas non-significant associations are shown in blue. The vertical dashed line represents the null effect (β = 0).

***Abbreviations:*** *act, cortical thickness; eTIV, estimated total intracranial volume; Rostral ACC, rostral anterior cingulate cortex; VentralDC, ventral diencephalon*

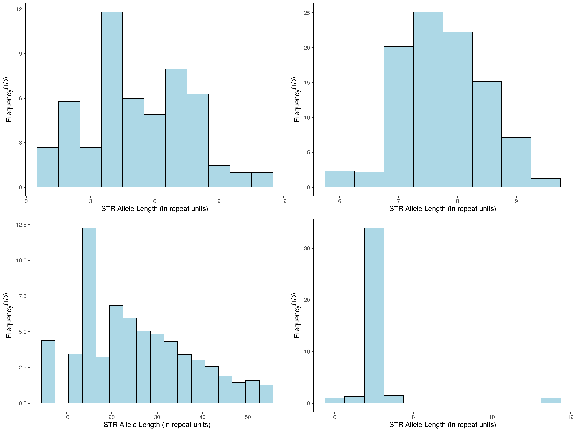

**Fig. S7. Distribution of allele sizes for *NADK* and *PRR14L* across both cohorts.** The x-axis denotes allele counts, and the y-axis shows the cube root of the allele frequency within each cohort. The top panel illustrates *NADK* distributions (RS on the left, UKB on the right), while the bottom panel shows *PRR14L* (RS on the left, UKB on the right). Allele distributions for *NADK* appear highly similar between cohorts, whereas *PRR14L* exhibits a broader range of allele sizes in RS compared to UKB, potentially reflecting population-level differences in repeat length variation or technical differences in sequencing read length

***Abbreviations:*** *NADK, Nicotinamide Adenine Dinucleotide Kinase; PRR14L, Proline-Rich Protein 14-Like*

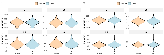

**Fig. S8. Distribution of STR expansion burden by sex across thresholds in both cohorts.**

Violin plots show the distribution of STR expansions per individual, stratified by sex (Female vs. Male), at defined expansion thresholds. Panels show Rhineland Study (A) and UKB (B). Across all thresholds, the distributions are similar between sexes, with median counts decreasing as the expansion threshold increases. These data indicate no significant sex differences in STR expansion burden in either cohort.

***Abbreviation:*** *UKB, UK Biobank*

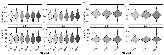

**Fig. S9. Distribution of polygenic STR expansion burden by age group across different thresholds in Rhineland Study and UKB.**

Violin plots display the distribution of STR expansion burden (y-axis) across age groups (x-axis) at increasing expansion thresholds. Panel A shows Rhineland Study (six age bins), and B panels show UKB (three age bins). In both cohorts, expansion burden remains largely stable across age groups, with only subtle, non-significant age-related increases observed at higher thresholds. The broader number of age bins in RS reflects its wider age range compared to UKB.

***Abbreviation:*** *UKB, UK Biobank*

**Table S1: Summary of Brain Imaging–Derived Phenotypes Across Cohorts**

| **Phenotype** | **Mean (SD) RS**  **n=2965** | **Mean (SD) UKB**  **n=38879** |
| --- | --- | --- |
| Accumbens | 4.82 (0.98) | 5.33(0.88) |
| Amygdala | 1.62 (0.22) | 1.88 (0.23) |
| Caudal ACC | 2.45 (0.14) | 2.42 (0.14) |
| Caudal Mid-Frontal | 2.47 (0.11) | 2.42 (0.12) |
| Caudate | 3.55 (0.48) | 3.45 (0.47) |
| Cerebelum cortex | 108.72 (12.08) | 111.3 (11.08) |
| Cerebral white matter | 458.21 (59.02) | 474.47 (56.69) |
| Cortical thickness | 2.45 (0.08) | 2.33 (0.08) |
| Cuneus | 1.95 (0.11) | 1.84 (0.1) |
| Entorhinal | 3.5 (0.23) | 3.16 (0.25) |
| Fusiform | 2.67 (0.1) | 2.52 (0.11) |
| Gray Matter | 627.36 (61.85) | 607.87 (52.98) |
| Hippocampus | 3.95 (0.45) | 4.17 (0.4) |
| Inf. Parietal | 2.42 (0.1) | 2.29 (0.11) |
| Inf. Temporal | 2.76 (0.11) | 2.64 (0.11) |
| Insula | 2.94 (0.13) | 2.95 (0.13) |
| Isthmus Cingulate | 2.3 (0.15) | 2.22 (0.13) |
| Lat. Occipital | 2.22 (0.11) | 2.07 (0.1) |
| Lat. Orbitofrontal | 2.54 (0.1) | 2.43 (0.1) |
| Lingual | 2.06 (0.1) | 1.91 (0.09) |
| Medial Orbitofrontal | 2.35 (0.13) | 2.33 (0.11) |
| Mid-Temporal | 2.75 (0.11) | 2.55 (0.11) |
| Pallidum | 1.96 (0.24) | 2.05 (0.25) |
| Paracentral | 2.46 (0.12) | 2.38 (0.14) |
| Parahippocampal | 2.77 (0.2) | 2.56 (0.23) |
| Pars Opercularis | 2.51 (0.11) | 2.43 (0.11) |
| Pars Orbitalis | 2.56 (0.13) | 2.44 (0.13) |
| Pars Triangularis | 2.34 (0.1) | 2.23 (0.11) |
| Post. Cingulate | 2.36 (0.11) | 2.32 (0.11) |
| Postcentral | 2.11 (0.11) | 1.98 (0.1) |
| Precentral | 2.58 (0.13) | 2.48 (0.14) |
| Precuneus | 2.37 (0.1) | 2.26 (0.12) |
| Putamen | 4.68 (0.60) | 4.68 (0.54) |
| Rostral ACC | 2.64 (0.14) | 2.58 (0.14) |
| SubCortGrayVol | 56.12 (5.88) | 56.93 (5.08) |
| Sup. Frontal | 2.52 (0.1) | 2.51 (0.12) |
| Sup. Parietal | 2.21 (0.1) | 2.07 (0.12) |
| Sup. Temporal | 2.83 (0.13) | 2.62 (0.13) |
| Supramarginal | 2.49 (0.11) | 2.39 (0.12) |
| Supratentorial | 976.55 (107.35) | 1058.42 (106.81) |
| TBV | 1112.21 (118.14) | 1108.83 (107.01) |
| Thalamus | 7.16 (0.91) | 6.63 (0.66) |
| Transverse Temporal | 2.41 (0.17) | 2.27 (0.19) |
| VentralDC | 396.8 (436.82) | 418.15 (416.26) |
| eTIV | 1551.50(148.10) | 1468.60(159.90) |

Table shows Mean (SD) values for brain imaging–derived phenotypes for the RS and UKB cohorts. Overall, distributions were comparable between the two cohorts.

***Abbreviations:*** *eTIV, estimated total intracranial volume; Rostral ACC, rostral anterior cingulate cortex; VentralDC, ventral diencephalon; RS, Rhineland Study; SubCortGrayVol, subcortical gray matter volume; TBV, total brain volume; UKB, UK Biobank. Volumes are in cm³; cortical thickness in mm.*

Table S2: Sequence parameters for the T1-weighted and T2-weighted imaging versions in the Rhineland Study

| **T1w sequence** | | | | **T2w sequence** | | | | |
| --- | --- | --- | --- | --- | --- | --- | --- | --- |
| Protocol | A multi-echo magnetization prepared rapid gradient echo (MPRAGE) sequence,^4^ with 2D acceleration.^5^ | | | Protocol | A 3D turbo-spin-echo (TSE) sequence, with variable flip angles.^6^ | | | |
|  | Version | | |  | Version | | | |
| Parameters | T1w^a^ | T1w^b^ | | Parameters | T2w^a^ | T2w^b^ | T2w^c^ | T2w^d^ |
| Repetion time (TR) | 2560 ms | | | Repetion time (TR) | 2800 ms | | | |
| Inversion time (TI) | 1100 ms | | | Echo time (TE) | 405 ms | | | |
| Matrix size | 320 × 320 × 224 | | | Matrix size | 320 × 320 × 224 | | | |
| Flip angle | 7° | |  | Phase-encoding direc.[^++^](javascript:;) | A>P | R>L | A>P | A>P |
| PI acc. factor | 1×3 | | 1×2 | PI acc. factor | 3×1 | | 2×1 | 1×2[^+^](javascript:;) |
| Echo time (TE) | 2.94 ms^*^ | | 1.68 ms to 6.51 ms^**^ | PI ref. scan | Integrated | | External | |
| Acquisition time (TA) | 3:43 minutes | | 6:35 minutes | Acquisition time (TA) | 3:57 minutes | 4:30 minutes | 4:47 minutes | |
| Readout bandwith | 240 Hz/pixel | | 740 Hz pixel |  |  |  |  | |

To date, there have been two versions of the T1w sequence (T1w^a-b^) and four versions of the T2w sequence (T2w^a-d^) - care was taken to preserve the image contrast between versions for both sequences.

^*^ 1 echo, ^**^ 4 echoes combined to 1.

^+^ with one CAIPIRINHA shift,^7^ ^++^ A: anterior, P: posterior, R: right, and L: Left.

Table S3.: Sequence parameters for the UK Biobank brain MRI protocol (31 minutes total scan time)

| **Modality** | **Duration** | **Voxel, Matrix** | **Key Parameters** |
| --- | --- | --- | --- |
| T1 | 4:54 | 1.0x1.0x1.0 mm  208x256x256 | 3D MPRAGE, sagittal, R=2, TI/TR=880/2000 ms |
| T2 FLAIR | 5:52 | 1.05x1.0x1.0 mm  192x256x256 | FLAIR, 3D SPACE, sagittal, R=2, PF 7/8, fat sat,  TI/TR=1800/5000 ms, elliptical |
| swMRI | 2:34 | 0.8x0.8x3.0 mm  256x288x48 | 3D GRE, axial, R=2, PF 7/8 TE1/TE2/TR=9.4/20/27  ms, |
| dMRI | 7:08 | 2.0x2.0x2.0 mm  104x104x72 | MB=3, R=1, fat sat, b=0(5x + 3x phase-encoding-  reversed), 1000(50x), 2000(50x) |
| rfMRI | 6:10 | 2.4x2.4x2.4 mm 88x88x64 | TE/TR=39/735 ms, MB=8, R=1, flip angle 52°, fat sat |
| tfMRI | 4:13 | 2.4x2.4x2.4 mm  88x88x64 | Acquisition same as rfMRI. Task is faces/shapes  “emotion” task. |

R = in-plane acceleration factor, MB = multiband factor, PF=partial Fourier. All non-EPI scans are pre-scan normalized (on-scanner bias-field corrected). Gradient distortion correction is turned off on the scanner and applied in post-processing.
